# Genetic *SNUPN* variants cause spinocerebellar atrophy by disrupting global splicing in Purkinje cells

**DOI:** 10.1101/2024.07.11.24310169

**Authors:** Mariko Okubo, Megumu Ogawa, Nobuyuki Eura, Yukiko U. Inoue, Ken-ichi Dewa, Tomohiro Owa, Satoshi Miyashita, Terumi Murakami, Hisayoshi Nakamura, Shinichiro Hayashi, Ikuya Nonaka, Katsuhisa Ogata, Mikio Hoshino, Takayoshi Inoue, Ichizo Nishino, Satoru Noguchi

## Abstract

We identified genetic variants in the *SNUPN* gene, which encodes the adapter protein snurportin-1 for the nuclear import of U1 snRNPs, in two families affected by spinocerebellar ataxia. We have elucidated the pathogenicity of these variants and the molecular pathomechanisms underlying this disease by assessing mutant snurportin-1 properties *in vitro*, cerebella at the morphological and molecular levels *ex vivo*, and motor functions in *Snupn*-variant knocked-in mice *in vivo*. Mutant snurportin-1 impaired nuclear-cytosol shuttling, leading to defective nuclear transport of U1 snRNPs in cerebellar Purkinje cells. This resulted in aberrant splicing and expression of genes essential for Purkinje cell development and impaired dendrite formation. The malformation of Purkinje cell dendrites resulted in hypoplasia and premature migration of granule cell precursors and interneurons, leading to abnormal lobe development and atrophy in the cerebellum.

---

Splicing is a mechanism that generates diverse gene products and determines spaciotemporal selectivity of the usage of alternative exons in genes. Pre-mRNA splicing is mediated by the spliceosome, a dynamic macromolecular complex that assembles anew on each intron^1^. Five small nuclear ribonucleoprotein particles (snRNPs) and hundreds of proteins participate in this process^2^. Among them, the U1 snRNP complex, which consists of U1 RNA and hetero-heptameric Sm ring proteins, initiates spliceosome assembly by binding to the 5’ splice site through base pairing between the 5’ terminal sequence of U1 RNA and a conserved stretch of nucleotides at the 5’ splice site of pre-mRNA. U1 snRNP also protects pre-mRNAs from premature cleavage and intronic polyadenylation^3-5^. Thus, U1 snRNP is essential for major splicing machinery and pre-mRNA stability with U2 snRNP. Recently, abnormalities in these splicing mechanisms are known to cause several neurodegenerative disorders.^6,7^

Nuclear transport of mature m3G-capped U1 snRNP through nuclear pore complexes is mediated by nuclear transport receptors, including importin beta. These receptors bind cargoes directly or through adapter molecules, constantly shuttling between the nucleus and cytoplasm by using the chemical potential of the nucleocytoplasmic RanGTP-gradient. For the transport of mature m3G-capped U snRNPs into nuclei, snurportin-1 is an import adapter for importin beta^8-10^. *In vitro* studies and structural analyses have characterized the functions of snurportin-1 well, whereas its physiological roles *in vivo* are not well documented.

Spinocerebellar atrophy (SCA) comprises a large group of heterogeneous neurogenerative disorders; however, it has common phenotypes such as cerebellar atrophy and ataxia, which are caused by the loss, immaturity, or degeneration of Purkinje cells (PCs). In recent years, many causative genes have been identified owing to the widespread clinical use of next-generation sequencing. To date, 49 types of SCAs have been characterized, and 38 causative genes have been identified^11-13^. In particular, recessive forms of SCA are congenital disorders, and several pathological mechanisms related to the mitochondrial respiratory chain complex, glutamic acid receptor, DNA repair, and autophagy have been reported^14-21^. Splicing abnormalities are also reported to cause some SCAs^6^. These abnormalities are frequently associated with PC physiology and dendrite malformation during cerebellar development^15,18-21^.

Here, we report genetic variations in the *SNUPN* gene, which encodes snurportin-1, in three patients from two SCA families. We generated pathogenic mouse models by mimicking the genetic variations in patients and showed that the mice recapitulated the phenotypes of human patients well. We also demonstrated decreased U1 snRNP nuclear transport and abnormal global gene splicing in PCs, which leads to defective dendrite formation, Shh secretion, and smaller cerebellar size due to decreased granule cell (GC) proliferation in the models. Thus, our study provides definitive evidence for the causative machinery of SCA, including splicing abnormalities in PCs, by dissecting the roles of snurportin-1 in cerebellar development.

## Results

### Patients with SCA in childhood

Three patients (from two families) exhibited cerebellar atrophy (especially in the vermis) but normal cerebral and medullary volumes (Fig. 1a). Their clinical symptoms are provided in Table 1. Patients 1 and 2 from Family A had apparent ataxia, whereas Patient 3 from Family B did not. All three patients had mild intellectual disability, and Patients 1 and 2 had congenital cataract. They showed different degrees of muscle weakness (Fig. S1a and Table 1).

**Fig. 1:**
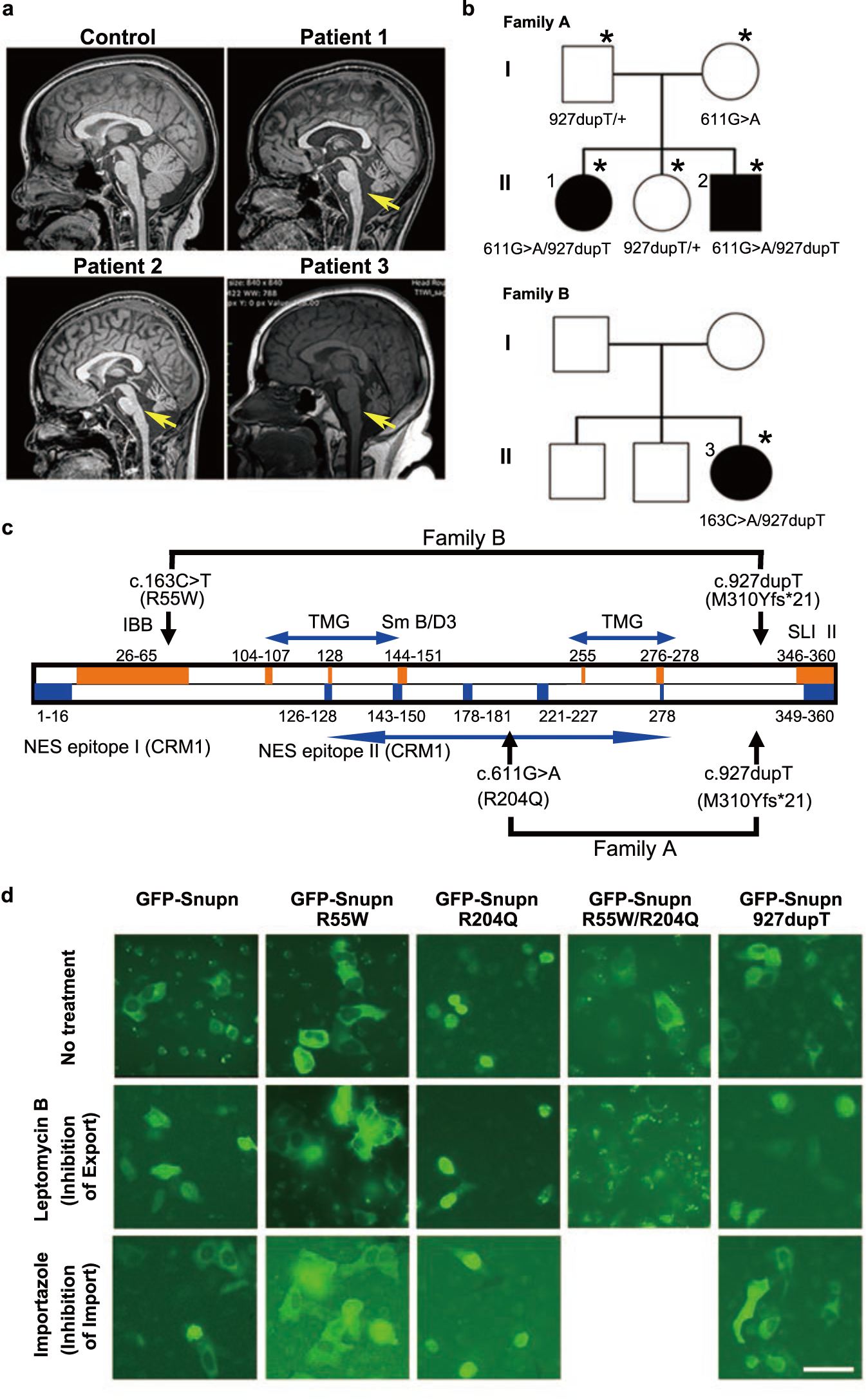
Characterization of *SNUPN* variants identified in patients. (a) T1-weighted sagittal midline images of a control individual (Control), Patient 1, Patient 2, and Patient 3.at age range of 10-15 years-old. Images of all patients show marked dilation of the cerebellar interfolia spaces and a marked decrease in size of the superior and inferior vermis (arrows). (b) The inheritance pattern of affected individuals is consistent with a fully penetrant autosomal recessive disorder. Squares and circles indicate males and females, respectively. Black-filled and open symbols indicate affected individuals a nd unaffected relatives, respectively. * denotes individual enforced whole exome sequencing (WES). (c) The location of identified snurportin-1 variants. Regions related to import/export into/from the nucleus are shown in orange and blue, r espectively. R55W is located at the binding domain with importinβ, and R204Q is located in the CRM1 binding domain. (d) Localization of wt and mutant GFP-snurportin-1 in HeLa cells after treatment with nuclear import and export inhibitors. Scale bar, 50 μm.

**Table 1.** Clinical information of patients.

|  | Patient 1 (A-II-1) | Patient 2 (A-II-3) | Patient 3 (B-II-3) |
| --- | --- | --- | --- |
| Age range at inspection | 11-15 | 6-10 | 11-15 |
| Ambulatory | Yes | No | No |
| Maintainance of standing position | Yes | Difficult | No |
| Ataxia | Mild | Severe | No |
| Ocular movement disorder | Yes | Yes | No |
| Dysarthria | No | No | No |
| Muscle weakness | Moderate<br>(Proximal > Distal) | Mild<br>(Proximal > Distal) | Severe<br>(Proximal > Distal) |
| Contraction | Ankle (6-10 y.o.) | No | No |
| Serum CK (IU/L) | 2336 (0-5 y.o.) | N/A | 801 |
| Cataract | Yes (0-5 y.o.) | Yes (0-5 y.o.) | No |
| Intellectual ability | IQ79 (11-15 y.o.) | IQ75 (6-10 y.o.) | Attend a special school |
| Age range at death (y.o.) | - | - | 11-15 |

(Age at inspection)

To identify the genetic cause of recessive cerebellar atrophy, we performed whole exome sequencing on two affected (II-1 and II-3) and three non-affected members (I-1, I-2, and II-2) in Family A and only one affected member (II-3) in Family B (Fig. 1b). The variants were filtered based on the assumption of autosomal recessive inheritance and with reference to the genetic databases of normal populations. Finally, we identified only three variants in the *SNUPN* gene: compound heterozygous c.611G>A and c.927dupT in Patients 1 and 2, and c.163C>T and c.927dupT in Patient 3 (Fig. S1b). The two missense mutations, c.611G>A and c.163C>T, were predicted to cause amino acid changes in snurportin-1, p.R204Q, and p.R55W and be “disease causing” by Polyphen2 (http://genetics.bwh.harvard.edu/pph2/) and Mutation Taster (https://www.mutationtaster.org/). A T duplication causes the truncation of the C-terminal 50 amino acid-sequence after methionine at 310. The location of these variants in the full-length molecule is shown in Fig. 1c.

### Pathogenicity of *SNUPN* variants in cultured cells

Localization of wild-type (wt) and mutant eGFP-snurportin-1 was observed 24 h after transfection to HeLa cells (Fig. 1d). Wt, R55W, and 927dupT-mutant eGFP-snurportin-1 were localized in the cytoplasm, whereas the R204Q-mutant was localized in nuclei. Snurportin-1 is known to shuttle between the cytoplasm and nucleus^22-25^. Thus, we used inhibitors of nuclear export (leptomycin B) and import systems (importazole). After treatment with leptomycin B, wt and 927dupT-mutants were localized in nuclei, indicating that these proteins were shuttled, whereas R55W remained in the cytoplasm. After treatment with importazole, R204Q-mutants were localized in nuclei, whereas R55W and R204Q double mutants were localized in the cytoplasm. These results indicate that R55W mutants cannot enter the nucleus, and that R204Q mutants cannot exit the nucleus.

Snurportin-1 has been shown to directly bind to importin and exportin during cytoplasmic nuclear transport. Thus, we analyzed the abilities of N-terminal peptides (1–65) of snurportin-1 to bind to importin beta by surface plasmon resonance. We found that binding between the N-terminal peptide of snurportin-1 and importin beta was weaker after R55W substitution (binding constant, KD_1_ = 3.9 × 10^-8^ M, KD_2_ = 1.0 × 10^-9^ M for wt; KD_1_ = 1.0 × 10^-7^ M, KD_2_ = 6.6 × 10^-8^ M for R55W; Fig. S2a). Conversely, we observed the fragmentation of the GFP-R55W/R204Q mutant 48 h after transfection (Fig. 1d). We also found decreased amounts of R204Q and R55W/R204Q mutations (Fig. S2b). We further analyzed the stability of the R204Q mutant by cycloheximide chase assays in HEK293 cells. Compared with wt and the other mutants, R204Q rapidly decreased (Fig. S2c), indicating its instability.

### R55W and I312Y* knock-in mice

We produced R55W and I312Y* knock-in mice using the CRISPR-Cas9 system. To characterize the variant-related phenotypes, we tried to generate homozygous mice by mating heterozygous female and male mice. For R55W, among the 26 young mice weaned, 7 were wt and 19 were heterozygous; however, no homozygous mice were obtained. We further analyzed embryonic stages 18.5, 13.5, and 8.5 days post-coitum; however, no homozygous embryos were identified. These results suggest that R55W might cause complete loss of snurportin-1 function. Conversely, I312Y* homozygous knock-in mice were obtained at a Mendelian rate and appeared normal.

To mimic the human patient’s genotypes, we generated compound heterozygotes harboring I312Y* and R55W. I312Y*/R55W was born at a Mendelian rate; however, approximately 40% of mice died by 3 weeks of age (Fig. 2a). After 3 weeks, the survival rate of the mice was similar to that of control I312Y*/+ littermates. However, both male and female I312Y*/R55W mice were smaller than I312Y*/+ mice (Fig. 2b, c). Behavioral tests were performed at 5 weeks of age. Gait was assessed with the footprint test, which showed that I312Y*/R55W mice had a wider hind-base width and shorter stride length than I312Y*/+ mice (Fig. 2d). The rotarod test revealed a significantly shorter riding period of I312Y*/R55W mice (Fig. 2e), which also exhibited a weaker grip strength (Fig. 2f). Interestingly, *ex vivo* gastrocnemius muscle force was also decreased; however, it was in the normal range when normalized by size (Fig. S3a–d). These results indicate that muscle weakness in I312Y*/R55W could be due to smaller muscle size (Fig. S3e). These mice also showed the limb clasping reflex that is associated with ataxia (Fig. 2g). Furthermore, the cerebella of I312Y*/R55W mice were significantly smaller in size and weight (Fig. 2h–j).

**Fig. 2:**
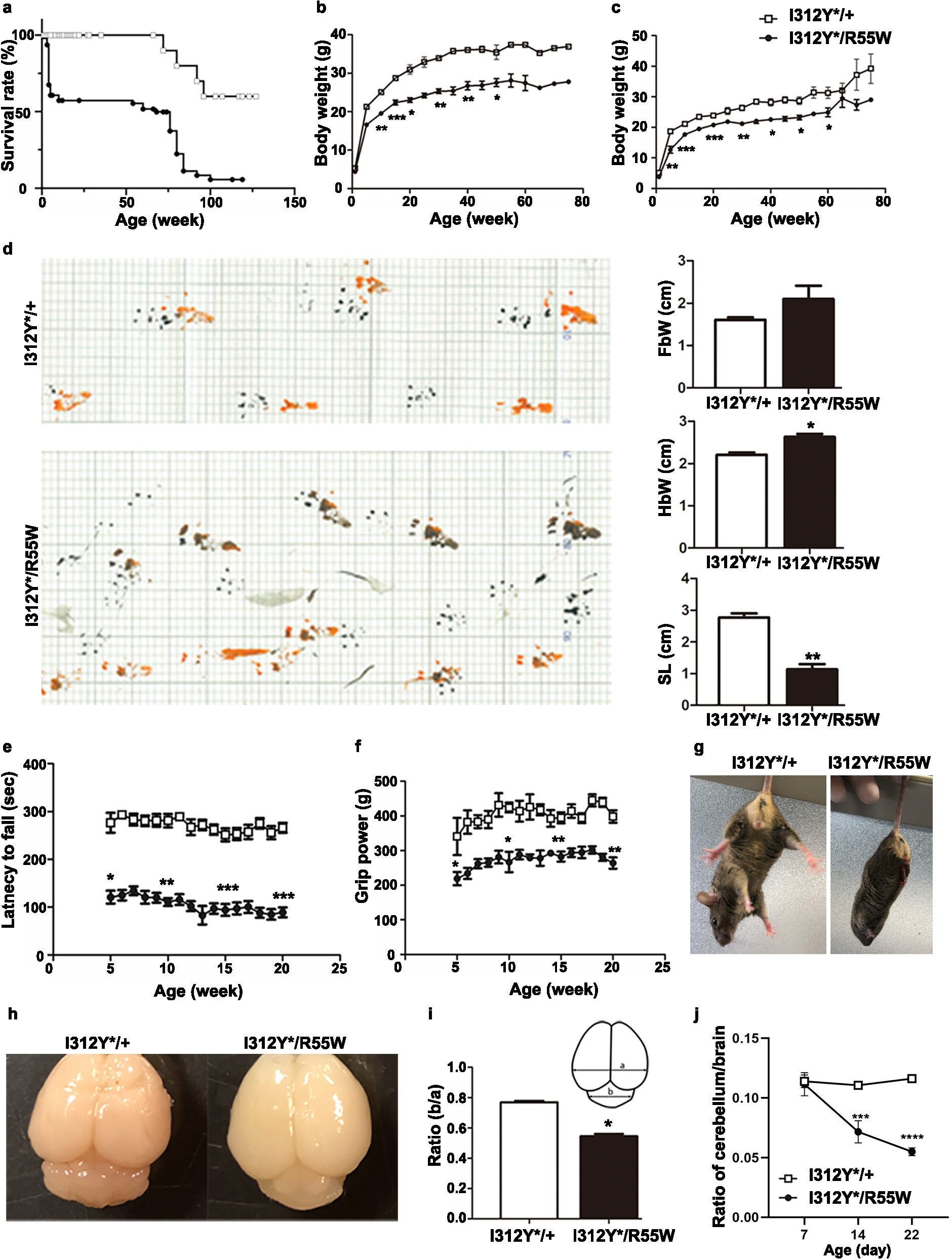
Physiological phenotypes of model mice. (a) Survival curves of I312Y*/R55W mice (black) and control I312Y*/+ littermates (white). (b) Growth (body weight) curves of male mice: I312Y*/+ (white, n = 3–12 per age group) and I312Y*/R55W (black, n = 4–6 per age group). (c) Growth (body weight) curves of female mice: I312Y*/+ (white, n = 4–12 per age group) and I312Y*/R55W (black, n = 2–12 per age group). (d) Gait of 18-month-old mice assessed by footprint assay. FbW: fore-base width, HbW: hind-base width, SL: Stride length (e) Time spent on a rotarod. I312Y*/+ (white, n = 5–12 per age group) and I312Y*/R55W (black, n = 5–12 per age group). (f) Grip power. I312Y*/+ (white, n = 5–12 per age group) and I312Y*/R55W (black, n = 5–12 per age group). (g) Limb clasping reflexes in I312Y*/+ and I312Y*/R55W mice. (h) Gross morphology of brains from adult I312Y*/+ and I312Y*/R55W mice. (i) The ratios (b/a) of transverse diameter of cerebrum and cerebellum. (j) Weights of cerebella during perinatal development. Data are presented as the mean ± SEM. *p < 0.05, **p < 0.01, ***p < 0.0001, ****p <0.00001.

### Morphological changes of PCs during cerebellar development in I312Y*/R55W mice

As I312Y*/R55W mice had smaller body weight, their skeletal muscles were smaller as well. Muscle examinations revealed no differences in muscle fiber numbers but smaller myofiber minFeret (minimum caliper diameter) in I312Y*/R55W than wt mice (Fig. S3g). However, no pathological changes were observed.

I312Y*/R55W mice represented cerebellar atrophy and exhibited growth retardation of cerebellum, as observed by different sizes in chronological sagittal sections of cerebella, but not regarding the structural architecture of lobes (Fig. 3a). Dendrite formation was poor in I312*/R55W P7 mice (Fig. 3b). At P24, Golgi staining showed poor dendrite development in I312Y*/R55W cerebella (Fig. 3c). Sholl analysis also revealed decreased arborization in I312Y*/R55W compared with I312Y*/+ mice (Fig. 3d). At P21, VGluT2-positive puncta were frequently observed at branched PC dendrites in I312Y*/+ mice, whereas VGluT2-puncta were observed near cell bodies in I312Y*/R55W mice (Fig. 3e–g), indicating that climbing fibers did not ascend distally along these dendrites. Also, at P14, we observed a similar distribution of VGluT2-puncta in I312Y*/R55W mice (data not shown).

**Fig. 3:**
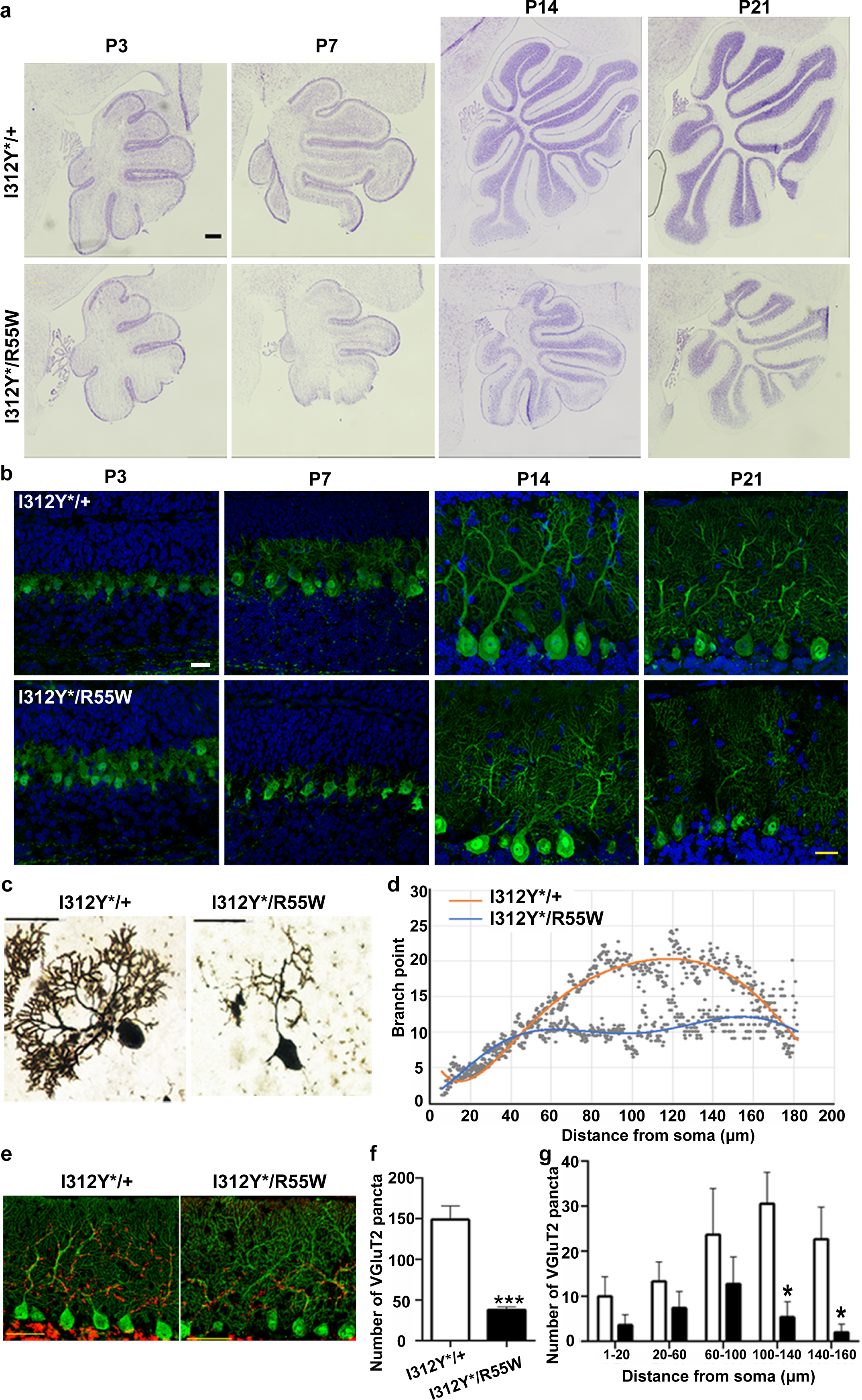
Morphological analyses of Purkinje cells (PCs) in model mice during perinatal development. (a) Nissl staining of sagittal cerebellar sections of I312Y*/R55W mice and littermate I312Y*/+ controls from P3 to P21. Scale bar, 200 μm. (b) Cerebellar PCs of control littermates (above) and I312Y*/R55W (below) from P3 to P21 labeled with calbindin-D28K (green) and DAPI (blue). Scale bars, 20 μm. (c) Golgi staining of PCs of I312Y*/+ and I312Y*/R55W mice at P24. Scale bars, 50 μm. (d) The number of branches revealed by Sholl analyses using Golgi staining of PCs. (e) Localization of VGluT2 (red) in PCs labeled with calbindin-D28K (green) of I312Y*/+ (left) and I312Y*/R55W mice (right) at P21. Scale bar, 100 μm. (f) The number of VGluT2 puncta in I312Y*/+ and I312Y*/R55W mice at P21. (g) The number of VGluT2 puncta at different distances from the PC soma at P21. Data are presented as the mean ± SEM. *p < 0.05, ***p < 0.0001.

### I312Y*/R55W mice exhibit decreased snRNP transport to PC nuclei

In cerebellar sections, snurportin-1 was strongly observed in the perinuclear surface of PCs in I312Y*/+ and I312Y*/R55W mice (Fig. 4a). In contrast to U1 snRNA, which was dispersed throughout the nucleus, small nuclear ribonucleoprotein D2 (SNRPD2), m3G-cap RNA, and survival motor neuron (SMN) proteins were concentrated and co-localized with coilin (Cajal bodies) in control I312Y*/+ mice (Fig. 4b, c). Interestingly, consistent with the impaired shuttling of mutant snurportin-1 in *in vitro* experiments, these snRNP proteins were hardly detected in Cajal bodies of PCs in I312Y*/R55W mice, indicating that nuclear transport of U1 snRNPs was impaired in mutant PCs. The distribution of U1 snRNA itself was not disturbed in I312Y*/R55W mice (Fig. 4d).

**Fig. 4:**
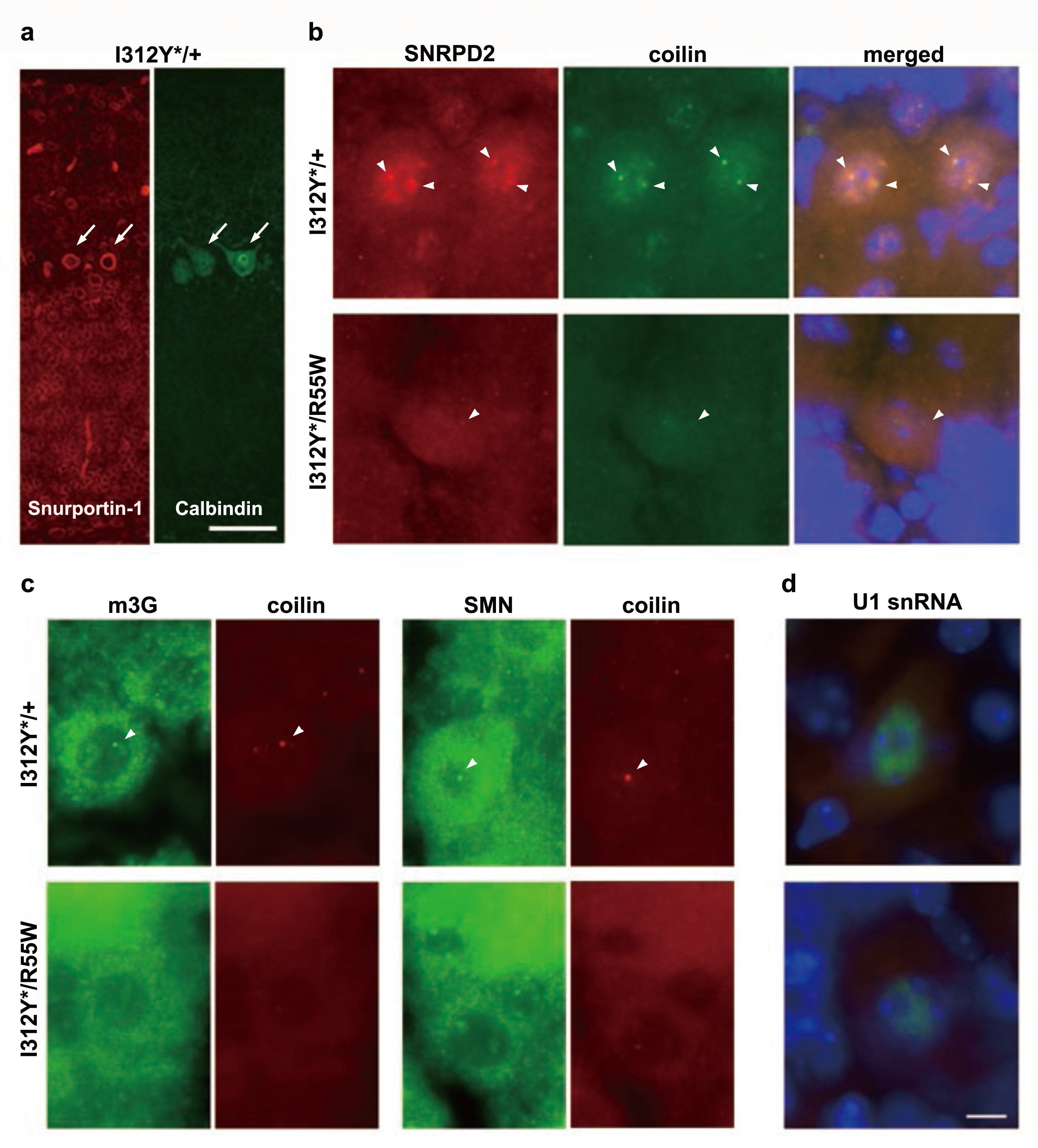
Defective transport of snRNPs into cerebellar Purkinje cell (PC) nuclei in model mice. (a) In mouse cerebellum, snurportin-1 was localized in the perinuclear region (red) in PCs, which were stained by calbindin antibody (green). Scale bar, 50 μm. (b), (c) U1 snRNP complex, SNRPD2 and m3G, and SMN proteins are less abundant in Cajal bodies (labeled with coilin) in cerebellar PCs from I312Y*/R55W mice. (d) Localization of U1 snRNA is not affected. Scale bar for (b), (c) and (d), 10 μm.

### Splicing abnormalities in genes expressed in PCs in I312Y*/R55W mice

To analyze splicing abnormalities in PCs, we identified 50 PC-specific genes from our single-cell RNA-sequencing (scRNA-seq) data and 365 PC-specific genes by searching the literatures (Table S1). Next, splicing abnormalities in these 415 genes in I312Y*/R55W mice were analyzed from whole cerebellum RNA-seq data using Mixture-of-Isoforms (MISO) software. We identified splicing changes in 134 genes (251 events), which showed more than 5% differences (Δψ > 0.05 or < −0.05), and classified them into five types (Fig. 5a and Table S2). We further confirmed the splicing changes in 11 genes by RT-PCR, indicating that the majority of aberrant splicing consisted of changes in alternative splicing (Fig. S4).

**Fig. 5:**
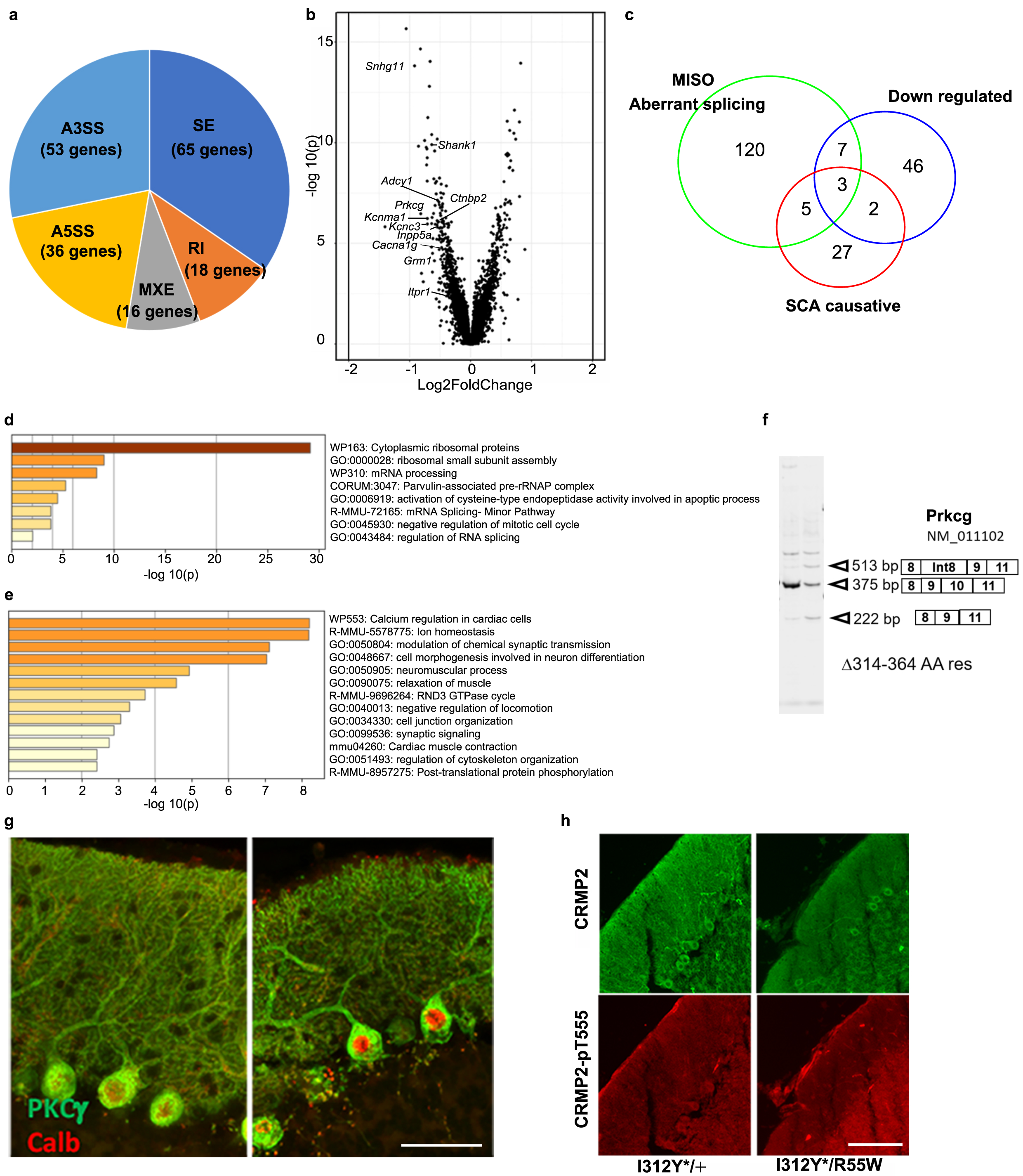
Altered splicing and gene expression in cerebellar Purkinje cells (PCs) in I312Y*/R55W mice. (a) The results of splicing alteration by MISO analysis. (b) Volcano plot of PC clusters with 52 upregulated and 57 downregulated genes (log fold change ≥ 0.5 or ≤ −0.5). (c) Venn diagram of genes with aberrant splicing, downregulated genes, and SCA-causative genes. (d), (e) Enriched ontology clusters of 52 upregulated genes (d) and 57 downregulated genes (e) by Metascape. (f) RT-PCR of Prkcg transcripts with exon 10 skipping or intron 8 insertion. (g) Localization of PKCgamma (green) in PCs labeled with calbindin (red) at P21 in control (left) and I312Y*/R55W (right) mouse cerebella. Scale bar, 100 μm. (h) Phosphorylation of PKC substrate, as observed with CRMP2 labeling in PCs. Staining with CRMP2 (above) and phospho-T555 CRMP2 (below). PCs in I312Y*/R55W mice were hardly stained with phospho-T555 CRMP2 antibody. Scale bar, 100 μm.

We identified 14,977 and 17,172 cells from I312Y*/+ and I312Y*/R55W cerebella at P6, respectively, and assigned distinct cellular clusters by specific cell lineage marker expression on Uniform Manifold Approximation and Projection (UMAP) (Fig. S5a, b). In both genotypes, PC clusters were composed of only 0.3% of all cerebellar cells (Fig. S5c). Morphological abnormalities in PCs were observed from P7 (Fig. 3a), indicating that PCs had the most extensive changes in gene expression among the different cell clusters observed by scRNA-seq. Differentially expressed genes between I312Y*/R55W and I312Y*/+ PCs were extracted on a volcano plot (Fig. 5b and Table S3). Interestingly, among the 58 downregulated genes, 10 genes were also included in aberrant spliced genes in I312Y*/R55W mice (Fig. 5c), suggesting that such splicing changes might eventually cause the reduction in the transcripts. Gene ontology enrichment analysis revealed that ion homeostasis, morphogenesis-related genes in neuronal development, and synaptic organization were enriched in the downregulated gene group, whereas protein synthesis, in particular, ribosomal proteins and snRNP-encoding genes were enriched in the upregulated group (Fig. 5d, e). Furthermore, five genes in the downregulated group were causative genes for SCA, of which three showed aberrant splicing. These results suggest that PC morphogenesis was affected at the transcript level in I312Y*/R55W mice. However, more interestingly, we could not find these up- and downregulated expression changes in nascent synthesized transcripts by single-nucleus RNA-seq (Fig. S6 and Table S4), indicating that these changes in gene expression might be associated with post-transcriptional events.

Furthermore, we analyzed the localization of *Prkcg* gene products, which showed the most drastic changes in aberrant splicing in I312Y*/R55W PCs. Mutations in this gene have been reported to be associated with SCA^13^. Representative aberrant splicing patterns for *Prkcg* are shown in Fig. 5f. The shorter product (222 bp) was produced by the skipping of exon 10, which is an in-frame exon spanning from the end of the C2 domain and N-terminal of the KD domain. Protein kinase C (PKC) gamma was highly and exclusively expressed in PCs and distributed in both cell bodies and dendrites in control mice (Fig. 5g). Its localization was similar in PCs in I312Y*/R55W mice; however, phosphorylation of the substrate, CRMP2 (pT155-CRMP2), was not detected (Fig. 5h). CRMP2 phosphorylation has been reported to regulate microtubule formation^26^, and we observed microtubules in the proximal domains of PC dendrites under electron microscopy. In control PCs, microtubules were well organized and associated with other cytoskeletal components, whereas in I312Y*/R55W PCs, microtubules were shorter and scattered, and only thinner filaments were organized (Fig. S7).

To explore the downstream events from changes in PCs, we analyzed sonic hedgehog (Shh) expression in PCs. scRNA-seq did not reveal any changes in Shh transcripts in PCs, whereas ELISA revealed significantly decreased cerebellar Shh protein levels (Fig. 6a, b). We also analyzed the consequence of Shh reduction in cerebellar scRNA-seq at P7. We classified proliferating GC progenitors (GCPs) and the differentiated GCs into five groups by UMAP clustering and drew pseudotime trajectories and heatmap (Fig. 6c, S8a and b). The numbers of proliferating GCPs were strikingly decreased, whereas those of differentiated GCs were increased in I312Y*/R55W mice (Fig. 6d). The numbers of GCPs were similarly decreased in all stages of the cell cycle. This suggests that there was no cell cycle arrest at any specific stage and that approximately one-quarter of cells withdrew from the cell cycle and differentiated into GCs. We did not find any significant differences in gene expression in the GCP or GC clusters between I312Y*/R55W and control mice (Table. S5), indicating that this is probably a non-cell autonomous phenomenon secondary to the reduction in Shh stimuli from PCs. We analyzed GCP proliferation by immunostaining I312Y*/+ and I312Y*/R55W cerebella with Mki67 at P7 (Fig. 6e, f). Interestingly, at the apical regions of lobes, stronger Mki67 staining was observed in external granule layer (EGL) in I312Y*/+ mice than in I312Y*/R55W mice, whereas Neurod1-positive cells were increased in the granular layer in I312Y*/R55W mice (Fig. 6g, i). However, basal regions of the lobes did not differ between the two mice (Fig. 6h, j).

**Fig. 6:**
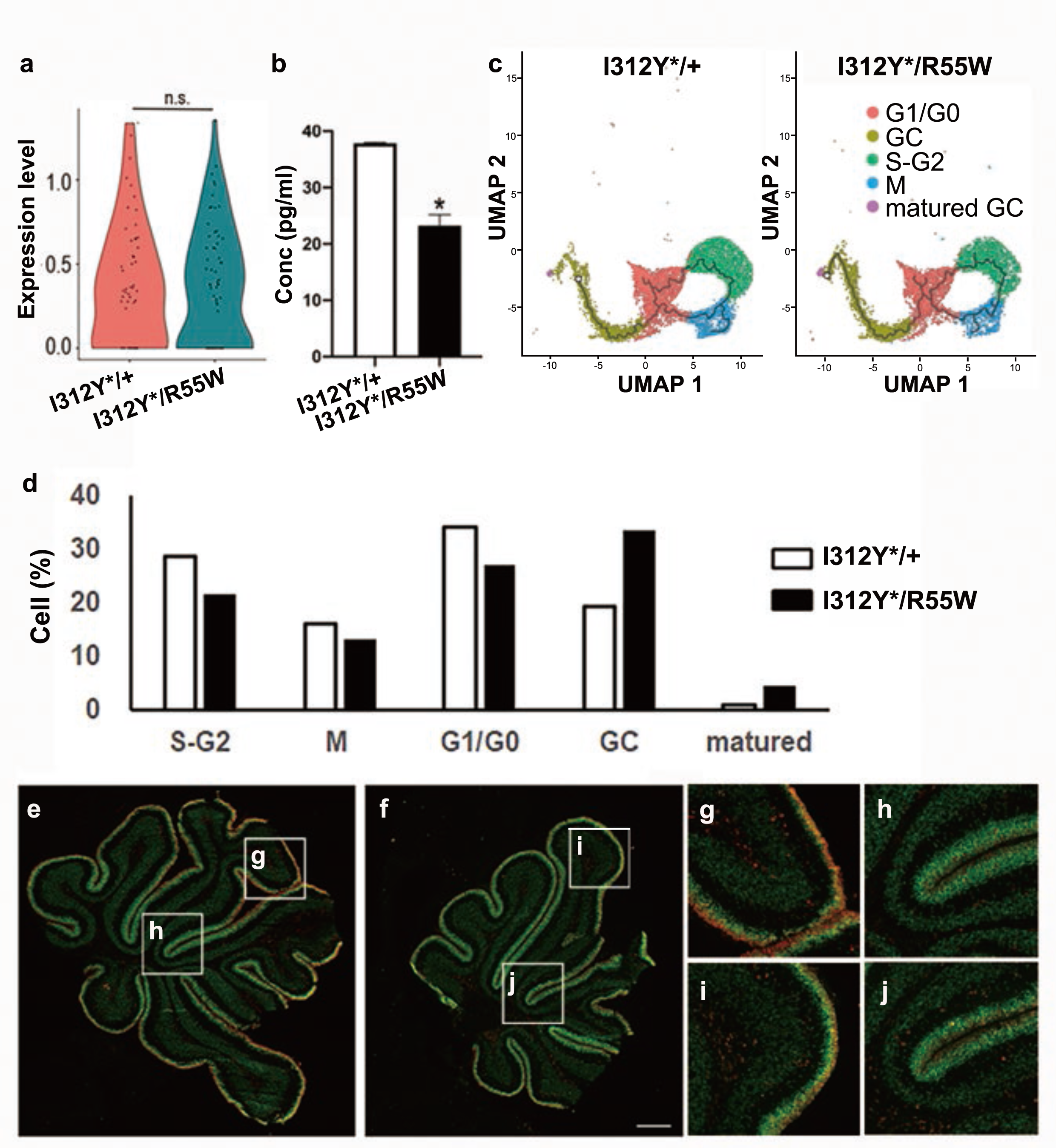
Decreased Shh secretion and withdrawal of granule cell progenitors (GCPs) from proliferation in I312Y*/R55W cerebella. (a) Expression of Shh transcript in Purkinje cells (PCs) and (b) Shh protein levels in cerebella (n = 6) from I312Y*/+ and I312Y*/R55W mice at P7. (c) Two-dimensional Uniform Manifold Approzimation and Projection (UMAP) distribution of cell clusters in the GC lineage of both mice. The GC lineage was divided into five clusters on the differentiation trajectory by R-analysis (resolution = 0.1). Each cluster was assigned according to specific gene expression (Fig. S8b). (d) Cell populations in the GC lineage. (e, f) Mki67-positive proliferating GCPs (red) and Neurod1-positive migrating GCs (green) in cerebella from I312Y*/+ (e) and I312Y*/R55W mice (f) at P7. Scale bar, 300 μm. At the apical regions of lobes, stronger Mki67 staining was observed in external granular layer (EGL) in I312Y*/+ (g) than that in I312Y*/R55W mice (i), whereas some Neurod1-positive cells migrated into the granular layer in I312Y*/R55W mice (i). Conversely, basal regions of lobes did not differ between the two mice (h, j).

We also found changes in inhibitory interneurons in I312Y*/R55W cerebella. According to gene expression, inhibitory interneurons were classified into four groups: Mki67-positive and Mki67-negative proliferating cells, ventricular zone-derived differentiated cells, and rhombic lip region-derived differentiated cells (Fig. S9a, b). In all clusters, inhibitory interneuron numbers were increased in I312Y*/R55W mice (Fig. S9c). We also detected increased Pax2-positive inhibitory interneuron numbers in the molecular layer and white matter in cerebellar sections from I312Y*/R55W mice (Fig. S9d).

## Discussion

In this study, we identified genetic variants of *SNUPN*, which encodes an adapter protein for nuclear import of U1 snRNPs, in two SCA families. We have clarified the pathogenicity of the variants and the underlying molecular mechanisms of SCA pathogenesis by studying the properties of mutant snurportin-1 *in vitro*, cerebella at the structural and molecular level *ex vivo*, and motor functions in Snupn-mutated mice *in vivo*. In brief, mutant snurportin-1 impairs nuclear-cytosol shuttling. It impairs U1 snRNP transport in cerebellar PCs, which leads to aberrant alternative splicing and defective dendrite formation in developing PCs. This decreases the proliferation and early maturation of developing GCs and inhibitory interneurons, which hinders lobe development in the cerebellum. This further results in abnormal synaptic networks with altered input to and output from PCs. (Fig. 7)

**Fig. 7:**
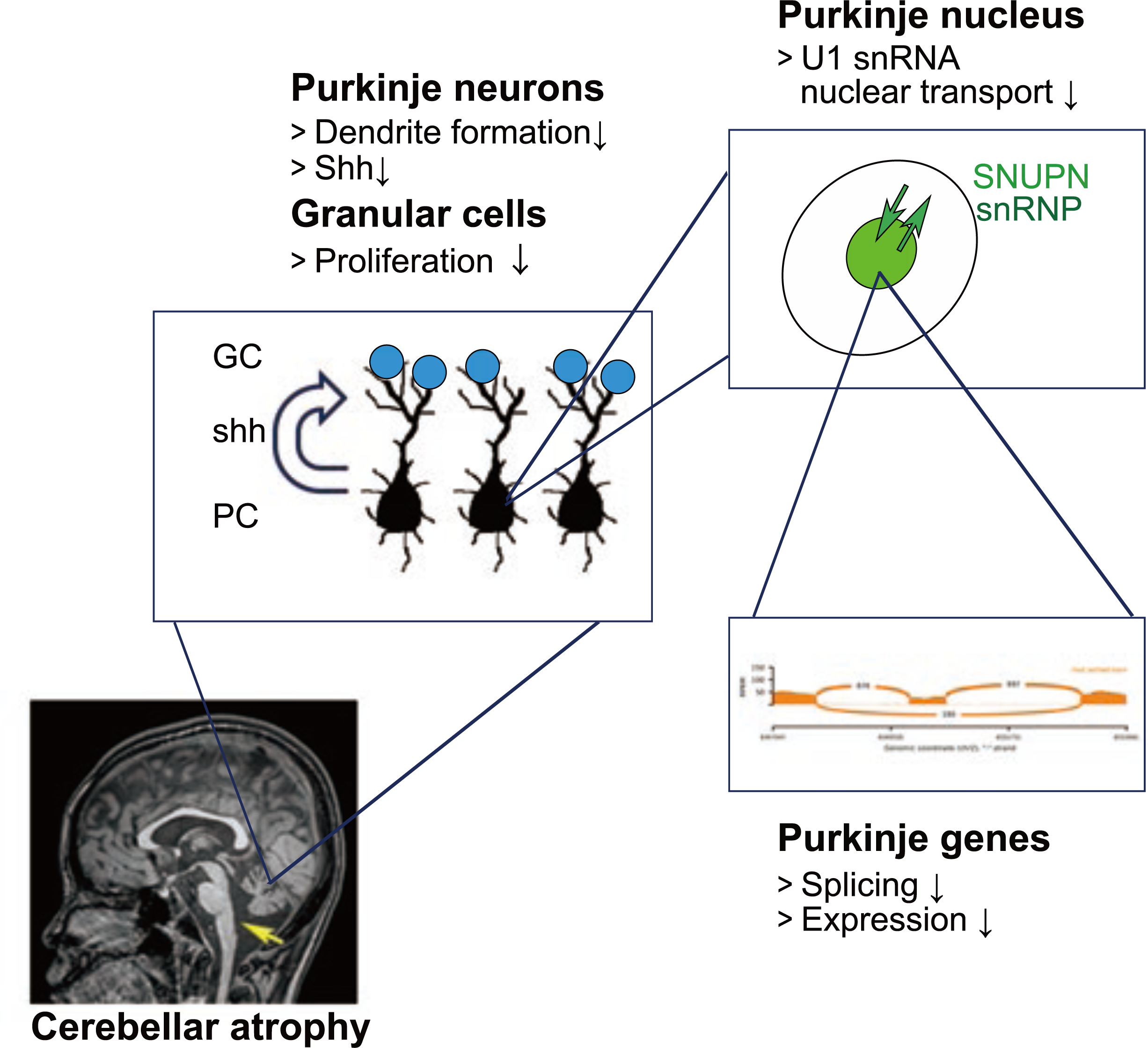
Conceptual diagram of disease caused by *SNUPN* mutations. Mutant snurportin-1 fails to shuttle between the nucleus and cytosol. This impaired shuttling decreases nuclear transport of U1 snRNPs in cerebellar Purkinje cells (PCs) and leads to global aberrant alternative splicing and decreased expression of genes essential for PC development. This global mis-splicing causes defective dendrite formation, which results in abnormal synaptic networks with altered input to and output from PCs. These PC defects cause hypoplasia and early maturation of developing granule cell (GC) progenitors and inhibitory interneurons. As a result, defects in the development of lobes and cerebellum occur.

We found three *SNUPN* variants in two SCA families. Two variants, c.611G>A and c.163C>T, were predicted to cause amino acid substitutions p.R204Q and p.R55W, respectively. Additionally, one common T duplication causes the truncation of 50 C-terminal amino acids. From the position of the variants in the functional map of molecular architecture^22, 23^ and our cell experiments, we concluded that the former missense amino acids are essential for nuclear-cytosolic transport of snurportin-1 and that their defects might affect several events during development, owing to the embryonic lethality of homozygous R55W mice. By contrast, C-terminal truncation might be milder but is essential for disease onset. The C-terminus of snurportin-1 has been reported to have dual functions by serving additional binding sites for stem-loop 3 of U1 snRNA in the import complex and exportin beta in the export complex^23, 27^. From observing such milder phenotypes, we speculated that such a truncated snurportin-1 might be functional; however, it decreased nuclear import and export shuttling, leading to decreased transport of U1 snRNPs into the nuclei of PCs. Additional studies to determine the precise function of truncated snurportin-1 are required.

Recently, two papers demonstrated pathogenic *SNUPN* variants that cause muscular dystrophy in humans^28,29^. Among the ten variants reported, nine of them were located at the C-terminus; however, none of these variants match those that we identified in our current study. Of note, one variant, c.164G>A, is predicted to be R55Q, which has a different amino acid substitution at the same residue as that of our variant. Interestingly, a patient who is homozygous for this variant has been reported to show a severe phenotype, which, however, does not include cerebellar atrophy^29^. In these papers, the main complaint of the patients harboring the *SNUPN* variants is limb girdle muscular dystrophy, even though some of the patients presented cerebellar atrophy. Conversely, muscle weakness of our patients was diverse (from mild to severe) but was predominantly observed in distal muscles in all patients. Furthermore, in our mouse model, muscle damage was not evident, and cerebellar atrophy was the main feature. We conclude that both limb girdle muscular dystrophy and SCA are allelic disorders caused by biallelic *SNUPN* variants. Further studies may provide the precise pathogenicity of each variant and detailed pathomechanisms underlying these different diseases.

The implications of splicing defects for SCAs have been broadly discussed. Several SCAs are caused by triplet repeat expansions^12,13^. This triplet expansion may cause diseases by disruptive splicing via cis effects on the expanded gene itself as well as trans effects on global aberrant splicing in many genes via sequestration of the splicing factors to nuclear foci by repeat expanded RNAs. More interestingly, mutations in genes related to U12 splicing, snRNA and its integral components, and splicing site binding proteins (*RNU12, RNPC3, ZRSR2, RNU4ATAC, STK11,* and *TRAPPC2*) are associated with several congenital diseases, including SCA^6^. Similarly, U2 snRNA gene (*Rnu2-8*) mutations were associated with neurodegeneration in mice that show global disruption and cerebellar atrophy. Interestingly, in these mice, not only changes in alternative 3’ splicing sites (which are recognized by U2 snRNP) but also global splicing changes (206 exons in 178 genes) in the whole cerebellum were observed. Recently, it has been reported that Alzheimer’s-disease-associated U1 snRNP splicing dysfunction causes neuronal hyperexcitability and cognitive impairment^30^. In this study, we identified 134 genes in PCs that were affected by splicing. This not only included recognition of the 5’ splicing site where U1 snRNP works, but also aberrant splicing as a major consequence. Interestingly, splicing was not completely affected, but only up to 30% in individual splicing events. These results indicate that numerous splicing events affected only to a small degree might be associated with disease onset. Thus, these findings might support previous insights into the pathogenesis of U2-related and repeat-expansion-related SCAs.

Using scRNA-seq, we identified downregulated genes related to ion homeostasis, morphogenesis during neuronal development, and synaptic organization, of which some showed aberrant splicing. We could not find these up- and downregulated changes by single-nucleus RNA-seq, indicating that these changes in gene expression might be associated with post-transcriptional events involved in regulating cytoplasmic pools of transcripts. Almost all the alternative splice transcripts we identified were in-frame splicing changes in transcripts with prevented nonsense-mediated RNA decays. However, we might have overlooked the splicing changes of downregulated genes with out-of-frame splicing changes. We also observed that ribosomal protein- and snRNP-encoding genes were upregulated in PCs. For the development of neurons and PCs, large amounts of proteins must be synthesized in the tips of dendrites or axons, and the ribosomal protein transcripts must be transported via axons/dendrites to the translated sites during early phases of PC development. However, dendritic defects may affect dendritic transport, resulting in protein accumulation in the soma. Similarly, the increase in snRNP-encoding genes might be a result of the decreased nuclear transport of snRNPs due to *Snupn* mutations in mice.

Many proteins and pathways participate in PC maturation and cerebellar development. Several papers have reported that impaired dendritic formation in PCs is tightly associated with SCA phenotypes. They involve mutations in the genes that encode cytoskeletal proteins (*MAP1A, MAP1B, MAP2,* and *TUBB3*) and signaling molecules (*PRKCG* and *CAMK2A*)^31-36^. Our findings regarding gene splicing, gene expression, and morphology suggest that malformation of PC dendrites plays a role in SCA pathogenesis. Notably, mutations in single genes affected the global splicing of numerous genes. Although the changes at the individual gene level were small, the cumulative defects of many genes contributed to impaired dendrite formation. This also suggests that mutant mis-spliced genes may cause disease.

Finally, our data suggest therapeutical applications, including supplementation of the *SNUPN* gene or even certain genes downstream of the more than 130 splicing-affected genes. For this, the associated critical pathological pathways must first be determined. However, as SCA is a congenital disorder due to developmental defects, this strategy might have some limitations. Nevertheless, by taking into consideration brain plasticity, the above-mentioned application for the correction of some mis-spliced genes might be useful to treat or modify SCA.

## Methods

### Ethic approval and consent to participate

All clinical information and materials used in the present study were obtained for diagnostic purposes with written informed consent. The study was approved by the Ethics Committee of National Center of Neurology and Psychiatry (Approved No. A2019-123, A2019-124). All animal care and experiments were performed in accordance with the Guidelines for Animal Experiments approved by the Animal Care and Use Committee of National Institute of Neuroscience, National Center of Neurology and Psychiatry (Approved No. 2022012).

### Whole exome sequencing

Whole exome sequencing was performed as previously described^37^. Genomic DNA was isolated from a muscle specimen of Patient 3 from Family B (Fig. 1b: B-II-3) and peripheral blood lymphocytes of two affected (Fig. 1b: A-Ⅱ-1, 2) and three non-affected members (Fig. 1b: A-Ⅰ and Ⅱ) from Family A. Genome annotation was performed by the National Center of Neurology and Psychiatry (NCNP) in-house pipeline as previously described^38^. Identified variants were confirmed by Sanger sequencing.

### Construction of plasmids expressing the snurportin-1 mutant

Wt human *SNUPN* ORF was amplified by PCR from the total RNA pool of the human brain. The expression vector pEGFP-*SNUPN* was constructed by cloning the amplified *SNUPN* ORF into the GFP-expression vector, pEGFP-C1, as described previously^39^. All snurportin-1 mutants were generated with the Site-Directed Mutagenesis Kit (Stratagene). Information on primer sequences is provided in Supplementary Table S6.

### Transfection of *SNUPN* constructs to HeLa cells and treatment with inhibitors of nuclear export and import

HeLa cells were cultured in Dulbecco’s Modified Eagle medium (DMEM; Thermo Fisher Scientific) supplemented with 10% fetal bovine serum (Life Technologies), penicillin (100 units/ml), and streptomycin (100 mg/ml). The expression vectors containing wt and mutant snurportin-1 were transfected using Lipofectamine with Plus Reagent (Thermo Fisher Scientific) into HeLa cells in Opti-MEM (Thermo Fisher Scientific). After 24 h, cells were treated with/without the nuclear export and import inhibitors leptomycin (10 ng/ml; Cayman) and importazole (40 μM; Abcam), respectively. After 1 h, localization of GFP-fluorescence was observed with a BZ-X700 microscope (KEYENCE).

To analyze mutant protein stability, the pEGFP-snurportin 1 constructs were transfected into HEK293T cells. After 24 h, cells were treated with 20 μg/ml cycloheximide. After 0, 1, 2, 4, 8, and 24 h, cells were harvested, and the GFP-snurportin-1 proteins were analyzed with western blotting using an anti-GFP antibody listed in Supplementary Table S7.

### Binding of the N-terminal fragment of snurportin-1 to importin beta

N-terminal fragments (1-65) of wt and R55W snurportin-1 were expressed as fusion protein with the C-terminal of glutathione S-transferase in *Escherichia coli*. Protein purification was performed as described previously. Binding of the snurportin-1 fusion proteins and recombinant human importin beta (Novus Biologicals) was measured by following a series of different importin beta concentrations (7.4, 22.2, 66.7, 200, and 600 nM) to the snurportin-1 fragments (2 μg/ml) fixed on a sensor chip with glutathione S-transferase antibodies. Surface plasmon resonance was measured by Wako Biochemicals.

### Design of CRISPR RNA (crRNA) and single-strand Oligo (ss-Oligo)

The crRNAs for production of R55W and I312Y* KI mice harboring the *Snupn* variant were designed by the CRISPR design tool (http://crispor.tefor.net/). The sequences of crRNAs for R55W and I312Y* along with trans-activating crRNA (tracrRNA) are shown in Supplementary Table S6. Single-stranded oligonucleotides, donors for homology-directed repair, were synthesized in Eurofins Genomics (Supplementary Table S6). Recombinant Cas9 (NEB, EnGen Cas9 NLS) was used for *in vitro* digestion assay on the amplified genomic fragments as previously described^40^.

### Microinjection of mouse on-cell embryos

KI mouse lines were generated via zygote electroporation utilizing the CRISPR/Cas9 system as described previousl^38, 41^. The founders of R55W and I312Y* KI mice were backcrossed on the C57BL/6J strain. A compound heterozygote (R55W/I312Y*) was generated by crossing heterozygous R55W KI mice with homozygous I312Y* mice. The heterozygous I312Y* KI littermates were asymptomatic and used as controls. The mice genotypes were identified by Sanger sequencing. Animals were kept in a barrier-free and specific pathogen-free grade facility on a 12 h light/12 h dark cycle and had free access to standard chow and water.

### Gate analyses

Body weight was measured weekly until 60 weeks (n = 3–12). Footprint tests were performed to measure stride length (SL), fore-base width (FbW), and hind-base width (HbW) at 18 weeks of age. The mice walked on an alley of 50 cm in length, 15 cm in width, and 15 cm in height. All their fore and hind paws were coated with nontoxic water-soluble ink (fore: black, hind: red), and the alley floor was covered with white paper. To obtain clearly visible footprints, at least three trials were conducted. FbW, HbW, and SL were measured manually.

### Rotarod test

Motor function was evaluated by a rotarod test consisting of a horizontal plastic rod with five divided lanes (Rota-Rod 47600: Ugo Basile). Control littermates, I312Y*/+ (n = 5–12 per age point), and I312Y*/R55W (n = 5–12 per age point) were measured at ages of 5–20 weeks.

Before the rotarod test, mice were trained by being forced to walk on the rods and at an accelerate speed of 2–40 rpm every 8 s for 5 consecutive days. For the test, mice were placed on a rotating rod at an accelerate speed of 2–40 rpm, and the latency to fall from the rod was recorded until a 300 s cut-off time. The mean latency to fall was calculated as the mean of three trials per mouse.

### Grip test

The grip strength of all four paws of 5–20-week-old mice was measured with a MK-380M grip strength meter (Muromachi Kikai) as described previously^42^. I312Y*/+ (n = 5–12 per age point) and I312Y*/R55W (n = 5–13 per age point) were measured. Grip strength was calculated as the mean of 10 trials per mouse.

### *Ex vivo* contractile and histological analysis of skeletal muscles

Mice were anesthetized by an intraperitoneal injection of midazolam, medetomidine hydrochloride, and butorphanol tartrate. The contractile forces of the tibialis anterior (TA) and gastrocnemius muscles were measured according to a previous protocol^42, 43^. I312Y*/+ and control wt (n = 3) were analyzed at 6 months of age. After the force measurements, the muscles were isolated, frozen at −160 ℃, and kept at −80 ℃ until use. Muscle sections were prepared with a cryostat. Hematoxylin and eosin and modified Gomori trichrome staining of muscle slices were performed as previously described^43^. The Minimum Feret diameter was measured according to previous protocols^43,44^.

### Cerebellum preparation

Mice were anesthetized and perfused with phosphate-buffered saline (PBS) and 4% paraformaldehyde (PFA). Then, isolated tissues were re-fixed in 4% PFA and cryoprotected with 30% sucrose in PBS. After embedding in optimal cutting temperature (OCT) compound, tissues were frozen in dry-ice-cold ethanol. For electron microscopy, mice were perfused with 4% PFA and 2% glutaraldehyde in PBS. For ELISA, mice were perfused with only PBS.

### ELISA

Cerebellum tissues of I312Y*/+ and I312Y*/R55W mice 7 days after birth were cut into 1–2 mm pieces and homogenized in PBS. ELISA was performed according to the protocol of the Mouse Shh N-Terminus Quantikine ELISA Kit (R&D systems).

### Immunohistochemistry and *in situ* hybridization of cerebellar sections

Cryosections of cerebellum 0, 3, 6, 7, 12 and 14 days after birth were cut with a thickness of 12–16 μm. Immunohistochemistry was performed according to a previous report^45^. Primary antibodies are described in Supplementary Table S7. Alexa-Fluor-conjugated donkey secondary antibodies (Invitrogen, 1:600) against species-specific IgGs corresponding to the primary antibodies were used. Fluorescence images were acquired using a BZ-X700 (KEYENCE) microscope. *In situ* hybridization for U1 snRNA was performed as described previously^46^. Probe information is shown in Supplementary Table S6.

### Golgi staining and Sholl analysis

The Golgi impregnation method for PC staining was performed according to the protocol of the FD Rapid GolgiStain Kit (FD Neuro Technologies). Cerebella of 28-day-old mice (n = 2, each of I312Y*/R55W and littermate controls) were stained. Thick sagittal sections of 100 μm were prepared in a cryostat. Images were acquired using a BZ-X700 (KEYENCE) microscope. Sholl analysis was performed on images using 10-μm spaced concentric circles (Sholl rings), and dendritic intersections were counted using ImageJ software. We analyzed representative PCs of I312Y*/+ (n = 8 PCs) and I312Y*/R55W mice (n = 6 PCs).

### Preparation of cells from cerebellum for scRNA-seq

Cerebella were separated from whole brains from P6 mice (6 days after birth) using tweezers, and the meninges were removed. Then the cerebella were sheared in 1×Hank’s balanced salt solution and digested with papain (10 μU/ml) for 30 min. Dissociated cells were filtered using 100 and 40 μm strainers, pelleted by centrifugation at 500×g for 5 min, re-suspended in Neurobasal medium (Thermo Fisher), and hemolyzed with Red Blood Cell (RBC) lysis buffer (QIAGEN). The residual cells were washed with PBS twice and finally collected by centrifugation. The cells were re-suspended in Neurobasal medium at a density of 1×10^6^ cells/ml. Cell viability was measured by trypan blue staining. For scRNA-seq, cell pools from cerebella of three mice for each genotype (I312Y*/+ and I312Y*/R55W) were analyzed. For RNA-seq, cerebella from six mice were independently analyzed.

### Preparation of nuclei of cerebellar cells for single-nucleus RNA sequencing (snRNA-seq)

Cerebella from one P6 I312Y*/R55W and one P6 I312Y*/+ mouse were separated from the whole brain using tweezers, and the meninges were removed. Cells were extracted according to the protocol of 10X GENOMICS Nuclei Isolation from Adult Mouse Brain Tissue for Single Cell RNA Sequencing (https://cdn.10xgenomics.com/image/upload/v1660261285/support-documents/CG000393_Demonstrated_Protocol_Adult_Mouse_Nuclei_Isolation_RevA.pdf). Dissociated cells were filtered using a 40-μm strainer. Then 7-amino-actinomycin D (7-AAD) was stained to remove dead cells using fluorescence-activated cell sortingFACS). Cell sorting was stopped at 300,000 cells because of the high cell count.

### Library construction and sequencing for scRNA-seq and snRNA-seq

The Chromium Next GEM Single Cell 3’ reagent kit (Dual Index) was used to construct the scRNA-seq and snRNA-seq libraries according to the manufacture’s protocol (10X Genomics). Briefly, isolated cells with nuclei were partitioned into nanoliter-scale Gel Bead-In-Emulsions (GEMs) in a limiting dilution to lower the multiplet rate. Then, single-cell GEMs were incubated with reverse-transcription reagents, including several primers for cell and transcript barcoding for cDNA synthesis, followed by cDNA amplification and library construction. Library sequencing was performed by BGI Japan.

### scRNA-seq and snRNA-seq data processing, filtering, and clustering

For scRNA-seq and snRNA-seq data processing, we used the Cell Ranger software provided by 10X Genomics, which helps convert 10X droplet data to metrics of expression counts. The data processing protocol has been described previously^47^. Cell distribution was visualized in a 2D t-distributed stochastic neighbor embedding (t-SNE) plot. We identified transcriptionally distinct cell clusters using the Seurat R package^48^.

### Differentially expressed gene analysis and trajectory analysis

The ‘FindAllMarkers’ function in Seurat and a threshold of 0.25 were used to identify unique cluster-specific marker genes, and ‘avg. logFC’ represents fold changes in the corresponding clusters. Violin plots, feature plots, and heart maps were generated using Seurat. The Monocle 3 package in R software program was used to determine the lineage of GCs.

### Analysis of RNA sequencing, expression, gene ontology, and alternative splicing

RNA was isolated with the RNeasy Mini Kit (QIAGEN) following manufacturer’s recommendations for RNA preparation from cells of whole cerebellum. All RNA samples were measured for quantity and quality. For sequencing, samples had to meet the minimum cut-off of 300 ng of RNA and an RNA integrity number (RIN) of more than 7. Library construction and sequencing was performed in BGI Japan. Briefly, library was constructed at the Broad Institute Genomics Platform using the same non-strand-specific protocol with poly-A selection of mRNA (Illumina TruSeq) used in the GTEx sequencing project to ensure consistency of our samples with GTEx control data. Paired-end 76 bp sequencing was performed on Illumina HiSeq 2000 instruments, with sequence coverage of 50 million or 100 million reads. Differentially expressed genes were identified using StrandNGS software (QIAGEN), and gene ontology was analyzed using Metascape software (https://metascape.org/gp/index.html#/main/step1).

To detect the alternative spliced genes from the RNA-seq data, MISO software was used^49^ (https://github.com/yarden/MISO/zipball/fastmiso). As PCs account for only 0.1% of total cerebellar cells, datasets were generated by combining RNA-seq data from four I312Y*/+ mice and five I312Y*/R55W mice. These two datasets were compared, and alternative events were extracted, including exon skipping (SE), alternative 3’/5’ splice site creation (A3SS, A5SS), mutually exclusive exon usage, (MXE) and intron retention (RI) by setting the cut-off value for differences as an absolute value of 0.05. Next, we aimed to clarify the primary splicing changes in PCs among the downstream aberrant splicing events in the other cell types. For this, we selected 415 genes, which were observed or reported to be especially expressed in PCs in mice for analysis (Supplementary Table S2) and analyzed aberrant splicing. The results were confirmed by RT-PCR using total RNAs from whole cerebella. Primers are listed in Supplementary Table S6.

## Data availability

The data that support the findings of this study are available from the corresponding author, S.N., upon reasonable request.

The RNA-seq datasets obtained in the current study are available in DDBJ Sequence Read Archive (DRA), accession number BioProject: PRJDB18388 (PSUB023433), available online.

## Code availability

No custom code was used for data analysis. All software and packages used are listed in the “Methods” section.

## Acknowledgements

We thank Ms. Yoko Tsutsumi for her technical assistance. Intramural Research Grant for Neurological and Psychiatric Disorders of NCNP (3-9, 5-6, and 5-7 to SN, MH, TI and IN) and by AMED under Grant Numbers JP23bm1223001s0102, JP23ek0109617s0402 (IN), JP23ak0101195s0101 (SN).

## Author contributions

S.N. and Ma.O. designed the study and developed conceptual ideas. Ma.O. performed *in vivo* experiments and analyzed all sequence data. Me.O. and S.N. performed *in vitro* experiments and abnormal splicing analyses. Y.U.I. and T.I. generated KI mice. N.E., S.M. and S.H. assisted single-cell/nucleus RNA-seq data analyses. K.D., T.O., M.H. and S.N. assessed brain anatomy and pathology, and performed immunostaining of mouse cerebella. H.N. performed electron microscopy. T.M., Ik.N, K.O., Ma.O. and Ic.N. participated in the clinical part of the study, muscle pathology, and genetic diagnosis of patients. Ma.O. and S.N. wrote the manuscript with suggestions from other authors.

## Competing interests

The authors have no relevant financial or non-financial interests to disclose.

## Additional Information

### Supplementary information

The online version contains Supplementary material available at…

## Additional information

**Supplementary information (Supplementary Figures 1–9 and Tables 1–7)**

## Notes

### Competing Interest Statement

The authors have declared no competing interest.

### Author Declarations

Ethic approval and consent to participate All clinical information and materials used in the present study were obtained for diagnostic purposes with written informed consent. The study was approved by the Ethics Committee of National Center of Neurology and Psychiatry (Approved No. A2019-123, A2019-124).

