## Supplementary information for "Genetic *SNUPN* variants cause spinocerebellar atrophy by disrupting global splicing in Purkinje cells"

**Table S1. List of genes specifically expressed in PC**

| Gene | NCBI Accession | Gene | NCBI Accession |
|---|---|---|---|
| *4930567K20Rik* | NR_046010.1 | *Kalrn* | XM_148399.5 |
| *4930587E11Rik* | NR_131017.1 | *Kcnab1* | NM_010597.2 |
| *9530059O14Rik* | NR_015610.2 | *Kcnd3* | NM_019931.1 |
| *9630028H03Rik* | NR_015544.2 | *Kcnh7* | NM_133207.1 |
| *A330015K06Rik* | NR_126080.1 | *Kcnip1* | NM_027398.2 |
| *Aars2* | NM_198608.2 | *Kcnj10* | NM_020269.3 |
| *Abat* | NM_172961.2 | *Kcnma1* | NM_010610.1 |
| *Abcd2* | NM_011994.1 | *Kcnq2* | NM_001003825.1 |
| *Abr* | NM_198018.1 | *Khdrbs2* | NM_133235.2 |
| *Adam23* | NM_011780.1 | *Kifc2* | NM_010630.1 |
| *Adamts4* | NM_172845.1 | *Kirrel2* | NM_172898.1 |
| *Adcy1* | mCT186681.0.0 | *Kitl* | NM_013598.1 |
| *Add3* | NM_013758.2 | *Klc2* | NM_008451.1 |
| *Adgrb3* | NM_175642.4 | *Ksr2* | NM_177751.2 |
| *Adrbk1 (Grk2)* | NM_130863.1 | *Large1* | NM_001317391.1 |
| *Actr3* | NM_023735.1 | *Lhfpl3* | XM_485569.1 |
| *Afap1l2* | NM_146102.1 | *Lingo2* | NM_175516.2 |
| *Aff1* | 5830443C17.1 | *Lpcat4* | NM_207206.1 |
| *Agbl4* | XM_485427.1 | *Lrch1* | XM_484393.1 |
| *Agmo* | NM_178767.2 | *Lrfn5* | NM_178714.2 |
| *Agtpbp1* | NM_023328.1 | *Lrp5* | NM_008513.1 |
| *Ahi1* | NM_026203.1 | *Lrp8* | NM_053073.1 |
| *Akap6* | XM_484140.1 | *Lrrfip1* | NM_008515.1 |
| *Aldh5a1* | NM_172532.1 | *Lrrtm4* | NM_178731.2 |
| *Ank2* | NM_178655.2 | *Macrod2* | XM_355346.2 |
| *Anks1b* | XM_618798.1 | *Magi2* | NM_015823.1 |
| *Ano10* | NM_133979.1 | *Maoa* | NM_173740.1 |
| *App* | NM_007471.2 | *Map1a* | XM_194040.4 |
| *Ar* | NM_013476.2 | *Map1b* | NM_008634.1 |
| *Arap2* | XM_132099.6 | *Map2* | NM_008632.1 |
| *Arhgap20* | NM_175535.3 | *Mapk1* | NM_011949.2 |
| *Arhgap26* | NM_175164.2 | *Mapk8ip2* | NM_021921.2 |
| *Arhgap31* | NM_020260.1 | *Mast4* | XM_283179.3 |
| *Arhgap32* | NM_177379.2 | *Mdga2* | NM_207010.1 |
| *Arhgap5* | NM_009706.1 | *Mdm4* | NM_008575.2 |
| *Arhgef33* | NM_001145452.1 | *Mef2c* | NM_025282.1 |
| *Astn2* | NM_019514.2 | *Meg3* | NM_144513.1 |
| *Atg4d* | NM_153583.8 | *Mettl16* | NM_026197.1 |
| *Atl2* | NM_019717.1 | *Mfsd11* | NM_178620.2 |
| *Atm* | NM_007499.1 | *Mthfr* | NM_010840.2 |
| *Atp2a3* | NM_016745.2 | *Mtpn* | NM_008098.2 |
| *Atp2b2* | NM_009723.1 | *Mtss1* | NM_144800.1 |
| *Atrnl1* | NM_181415.4 | *Muc3* | XM_355711.2 |
| *Atxn1* | NM_009124.2 | *Myo10* | NM_019472.1 |
| *Atxn2* | TC1461838.1 | *Napb* | NM_019632.1 |
| *Auts2* | NM_001363480.1 | *Nbea* | TC1429975.1 |
| *Axin1* | NM_009733.1 | *Ncam1* | NM_010875.2 |
| *Bax* | NM_007527.2 | *Ncoa1* | NM_010881.1 |
| *Bcl11a* | NM_016707.1 | *Ndnf* | NM_172399.1 |
| *Bcl2* | NM_009741.2 | *Ndrg4* | NM_145602.1 |
| *Bdnf* | NM_007540.3 | *Ndst1* | NM_008306.2 |
| *Bzrap1* | NM_172449.1 | *Ndufa5* | NM_026614.1 |
| *Cacna1a* | NM_007578.1 | *Nell1* | NM_177413.2 |
| *Cacna1b* | NM_007579.1 | *Neurog1* | NM_010896.2 |

| | | | |
|---|---|---|---|
| *Cacna1c* | NM_009781.1 | *Neurog2* | NM_009718.2 |
| *Cacna1d* | NM_028981.1 | *Ngfr* | NM_033217.1 |
| *Cacna1g* | NM_009783.1 | *Nktr* | NM_010918.1 |
| *Cacna1s* | XM_358335.2 | *Nlgn1* | NM_138666.2 |
| *Cacna2d2* | NM_020263.2 | *Nlgn2* | NM_198862.2 |
| *Cacnb4* | TC1462712.1 | *Nmdar* | NM_001372558.1 |
| *Calb1* | TC1568100.1 | *Nova2* | NM_001029877.3 |
| *Calb2* | NM_007586.1 | *Nptxr* | NM_028763.2 |
| *Camk2b* | NM_007595.2 | *Nrcam* | NM_176930.2 |
| *Camk2n2* | NM_028420.2 | *Nrk* | NM_013724.1 |
| *Camk4* | NM_009793.1 | *Nrp2* | NM_010939.1 |
| *Camta1* | XM_355539.2 | *Nrxn3* | NM_172544.1 |
| *Car8* | NM_007592.1 | *Nsd1* | NM_008739.1 |
| *Casc4* | NM_177054.3 | *Ntrk2* | NM_008745.1 |
| *Cask* | NM_009806.1 | *Ntrk3* | NM_008746.4 |
| *Ccdc64 (Bicdl1)* | XM_132372.3 | *Nup93* | NM_172410.1 |
| *Ccdc85a* | NM_181577.2 | *Nwd2* | XM_132047.4 |
| *Ccdc88a* | NM_176841.2 | *Olig2* | NM_016967.1 |
| *Cck* | NM_031161.1 | *Olig3* | NM_053008.1 |
| *Cd38* | NM_007646.2 | *Omp* | NM_011010.2 |
| *Cdc42* | NM_009861.1 | *Oxsr1* | XM_135264.5 |
| *Cdh12* | NM_001008420.1 | *P2rx4* | NM_011026.1 |
| *Cdh18* | XM_354862.2 | *Paxbp1* | XM_358888.2 |
| *Cdk14* | NM_011074.1 | *Pbrm1* | NM_025847.1 |
| *Celf2* | NM_010160.1 | *Pcdh19* | XM_205287.3 |
| *Cend1* | NM_021316.2 | *Pcp2* | NM_008790.1 |
| *Cep126* | NM_001045524.2 | *Pcp4* | NM_008791.1 |
| *Chd5* | XM_196334.4 | *Pcsk6* | XM_355911.2 |
| *Chgb* | NM_007694.1 | *Pde5a* | NM_153422.1 |
| *Clmn* | NM_053155.1 | *Pde9a* | NM_008804.2 |
| *Clstn3* | NM_153508.2 | *Per1* | NM_011065.2 |
| *Cntnap1* | NM_016782.1 | *Phactr2* | XM_125520.5 |
| *Cntn1* | NM_007727.1 | *Pkp4* | NM_026361.1 |
| *Cntn3* | NM_008779.1 | *Plcb1* | NM_019677.1 |
| *Cntn5* | XM_146640.5 | *Plcb4* | NM_013829.1 |
| *Cntnap2* | NM_001004357.0 | *Plcxd2* | XM_484546.1 |
| *Cntnap5a* | NM_001371013.1 | *Plekhd1* | XM_126991.4 |
| *Cntnap5b* | NM_172851.1 | *Pmpca* | NM_173180.2 |
| *Cntnap5c* | NM_001081653.2 | *Poldip3* | NM_178627.1 |
| *Col18a1* | NM_009929.2 | *Polrmt* | NM_172551.1 |
| *Corin* | NM_016869.1 | *Ppp1r16b* | NM_153089.2 |
| *Cpeb1* | NM_007755.1 | *Ppp1r17* | NM_011153.2 |
| *Cpne1* | NM_170590.1 | *Ppp1r1b* | NM_144828.1 |
| *Cpne9* | NM_170673.3 | *Ppp4r4* | NM_028980.1 |
| *Cpsf1* | NM_053193.1 | *Prex1* | XM_485102.1 |
| *Crhr1* | NM_007762.2 | *Prex2* | XM_129358.3 |
| *Csdc2* | NM_145473.1 | *Prkcg* | NM_011102.2 |
| *Csde1* | NM_144901.2 | *Prkg1* | NM_011160.1 |
| *Cttnbp2* | XM_289703.3 | *Prpf38b* | NM_025845.1 |
| *Ctxn2* | C630031J20.1 | *Ptbp1* | NM_008956.1 |
| *Cwf19l1* | XM_129328.5 | *Ptchd4* | XM_140020.3 |
| *Dab1* | NM_010014.1 | *Pten* | NM_008960.2 |
| *Dach1* | NM_007826.1 | *Ptf1a* | NM_018809.1 |
| *Ddx26b* | NM_172779.1 | *Ptprm* | NM_008984.1 |
| *Ddx55* | NM_026409.4 | *Ptprz1* | XM_358362.2 |
| *Dgkb* | NM_178681.2 | *Pura* | NM_008989.4 |

| | | | |
|---|---|---|---|
| *Dgkg* | NM_138650.1 | *Pvalb* | NM_013645.2 |
| *Dgkh* | XM_484397.1 | *Rabggtb* | NM_011231.1 |
| *Dgki* | XM_355752.2 | *Rasal2* | XM_355247.3 |
| *Dgkz* | NM_138306.1 | *Rb1* | NM_009029.1 |
| *Disp2* | NM_170593.1 | *Rbfox1* | NM_021477.2 |
| *Dkk3* | NM_015814.2 | *Rbfox2* | NM_053104.3 |
| *Dlg2* | NM_011807.1 | *Rbms1* | NM_020296.1 |
| *Dmd* | NM_007868.1 | *Rbms3* | NM_178660.2 |
| *Dmxl2* | XM_358382.2 | *Rell2* | NM_153793.2 |
| *Dner* | NM_152915.1 | *Reln* | NM_011261.1 |
| *Dpp10* | NM_199021.2 | *Rgs7bp* | NM_029879.1 |
| *Dscam* | NM_031174.2 | *Rgs8* | NM_026380.2 |
| *Ebf1* | NM_007897.1 | *Rictor* | NM_030168.2 |
| *Ebf2* | NM_010095.2 | *Rims2* | NM_053271.1 |
| *Efr3a* | NM_133766.1 | *Robo2* | NM_175549.2 |
| *Egfem1* | XM_130829.4 | *Rora* | NM_013646.1 |
| *Elmo1* | NM_080288.1 | *Rreb1* | XM_127279.5 |
| *Emd* | NM_007927.1 | *Ryr1* | NM_009109.1 |
| *Eml6* | NM_146016.1 | *Sacs* | NM_172809.1 |
| *Enho* | NM_027147.1 | *Sam68 (Khdrbs1)* | NM_011317.2 |
| *Epb4.1l3* | NM_013813.1 | *Scg3* | NM_009130.1 |
| *Epha4* | NM_007936.2 | *Scn1a* | XM_619757.1 |
| *Epha5* | NM_007937.1 | *Scn1b* | NM_011322.2 |
| *Epha6* | NM_007938.1 | *Scn8a* | NM_011323.1 |
| *Erc2* | NM_177814.3 | *Sema3a* | NM_009152.2 |
| *Ercc6l* | NM_146235.2 | *Setx* | NM_198033.1 |
| *Etl4* | TC1571230.1 | *Sez6l2* | NM_144926.2 |
| *Exosc8* | NM_027148.2 | *Sf3a1* | NM_026175.3 |
| *Fam107b* | NM_025626.3 | *Sf3b1* | NM_031179.1 |
| *Fam149b* | NM_172379.1 | *Sgcz* | NM_145841.1 |
| *Fam184b* | NM_021416.2 | *Shank1* | NM_001034115.1 |
| *Far2* | NM_178797.2 | *Shank2* | XM_146170.3 |
| *Fbxo33* | XM_127032.3 | *Shh* | NM_009170.2 |
| *Fcor* | XM_358687.2 | *Shisa6* | XM_354612.2 |
| *Fmr1* | NM_008031.2 | *Slc12a5* | NM_020333.1 |
| *Foxp1* | NM_053202.1 | *Slc17a8* | NM_182959.2 |
| *Foxp2* | NM_053242.3 | *Slc1a3* | NM_148938.2 |
| *Foxp4* | NM_028767.1 | *Slc1a6* | NM_009200.1 |
| *Frmd4a* | NM_172475.2 | *Slc35f1* | NM_178675.3 |
| *Frmd5* | NM_172673.2 | *Slc4a10* | NM_033552.1 |
| *Frmpd3* | NM_177750.2 | *Slc6a8* | NM_133987.1 |
| *Frmpd4* | XM_285569.3 | *Slc9a1* | NM_016981.1 |
| *Fsd1* | NM_183178.1 | *Slc9a3* | XM_127434.4 |
| *Fubp1* | NM_057172.1 | *Slc9a6* | NM_172780.1 |
| *Fxyd7* | NM_022007.1 | *Snap25* | NM_011428.2 |
| *Gabra1* | NM_010250.2 | *Sncb* | NM_033610.2 |
| *Gabrb2* | NM_172432.1 | *Snhg11* | NM_175692.2 |
| *Gabrb3* | NM_008071.2 | *Sntg1* | NM_027671.3 |
| *Gabrg2* | NM_008073.1 | *Snx14* | NM_172926.1 |
| *Galnt13* | NM_173030.1 | *Sos1* | NM_009231.1 |
| *Garnl3* | NM_178888.3 | *Sox5* | NM_011444.1 |
| *Gdnf* | NM_010275.1 | *Spock3* | NM_023689.2 |
| *Gfod1* | XM_488675.1 | *Sptan1* | XM_207079.3 |
| *Gfra1* | NM_010279.2 | *Sptbn2* | NM_021287.2 |
| *Gm14033* | ENSMUST00000128307 | *Srgap1* | XM_125904.3 |
| *Gm15336* | ENSMUST00000128894 | *Stac* | NM_016853.1 |

| | |
|---|---|
| *Gm26713* | NSMUST00000181827. |
| *Gm32647* | NSMUST00000206116. |
| *Gm35188* | NSMUST00000218869 |
| *Gm42439* | NSMUST00000195931 |
| *Gm44040* | NSMUST00000203360 |
| *Gm44071* | NSMUST00000204180 |
| *Gng13* | NM_022422.3 |
| *Gng4* | NM_010317.2 |
| *Gnptab* | NM_001004164.1 |
| *Gpc6* | NM_011821.1 |
| *Gria2* | NM_013540.1 |
| *Gria3* | NM_016886.1 |
| *Gria4* | NM_019691.2 |
| *Grid2* | NM_008167.1 |
| *Grid2ip* | NM_001159321.2 |
| *Grik1* | NM_146072.1 |
| *Grin1* | NM_008169.1 |
| *Grip1* | NM_028736.1 |
| *Grm1* | NM_016976.1 |
| *Grm5* | XM_149971.4 |
| *Grm7* | XM_144986.3 |
| *Gstm1* | NM_010358.2 |
| *Hdac9* | NM_024124.2 |
| *Heatr5a* | NM_177171.3 |
| *Hepacam* | NM_175189.2 |
| *Homer3* | NM_011984.1 |
| *Hpcal1* | NM_016677.1 |
| *Hsd11b1* | NM_008288.1 |
| *Hspa12a* | NM_175199.1 |
| *Hspa14* | NM_015765.1 |
| *Htr1b* | NM_010482.1 |
| *Htt* | NM_010414.1 |
| *Huwe1* | XM_136108.4 |
| *Iltifb* | NM_054079.2 |
| *Immp2l* | NM_053122.2 |
| *Inpp1* | NM_008384.1 |
| *Inpp4a* | NM_030266.1 |
| *Inpp5a* | NM_183144.1 |
| *IREB2* | XM_134910.4 |
| *Itgb3* | NM_016780.1 |
| *Itpka* | NM_146125.1 |
| *Itpr1* | NM_010585.5 |

| | |
|---|---|
| *Stam* | NM_011484.2 |
| *Stat3* | NM_011486.3 |
| *Stk39* | NM_016866.2 |
| *Stmn3* | NM_009133.2 |
| *Strn3* | NM_052973.1 |
| *Stx3* | NM_011502.1 |
| *Sugp2* | NM_172755.2 |
| *Svil* | NM_153153.1 |
| *Sycp1* | NM_011516.1 |
| *Syn1* | NM_013680.1 |
| *Syndig1* | XM_485077.1 |
| *Syt14* | NM_181546.2 |
| *Tbl1xr1* | NM_030732.3 |
| *Tbp* | NM_013684.1 |
| *Tbpl1* | NM_011603.2 |
| *Tenm2* | NM_011856.2 |
| *Tenm3* | NM_011857.2 |
| *Tenm4* | NM_011858.1 |
| *Thsd7a* | XM_287555.3 |
| *Thy1* | NM_009382.2 |
| *Tpp1* | NM_009906.2 |
| *Trabd2b* | NM_001085549.1 |
| *Traf6* | NM_009424.2 |
| *Trim32* | NM_053084.1 |
| *Trim9* | NM_053167.1 |
| *Trpc3* | NM_019510.1 |
| *Ttll5* | XM_126935.5 |
| *Ube3a* | NM_011668.1 |
| *Ube3b* | NM_054093.1 |
| *Unc5b* | NM_029770.1 |
| *Unc5d* | NM_153135.1 |
| *Upf1* | NM_030680.1 |
| *Vav3* | NM_020505.1 |
| *Vps13d* | XM_204062.4 |
| *Vstm2l* | NM_198627.2 |
| *Washc2* | NM_026585.3 |
| *Wnt3* | NM_009521.1 |
| *Wwox* | NM_019573.2 |
| *Zdhhc14* | NM_146073.3 |
| *Zfand4* | XM_132758.5 |
| *Zfp385c* | NM_177790.2 |

**Table S2. Genes with splicing abnormalities in PC by MISO analysis**

| Gene | NCBI Accession (cerebellum transcript) | Splicing pattern | delta Ψ | Event location |
|---|---|---|---|---|
| *Abat* | NM_172961.3 | SE | -0.06 | chr16:8608856:8608993:chr16:8611127:8611294:chr16:8613983:8614129 |
| *Abat* | NM_172961.3 | A5SS | 0.2 | chr16:8582737:8582782\|8582854:chr16:8588886:8588935 |
| *Adam23* | NM_011780.3 | SE | -0.07 | chr1:63572887:63572998:chr1:63585365:63585455:chr1:63592608:63596515 |
| *Adcy1* | | SE | 0.09 | chr11:7165412:7165526:chr11:7166240:7166289:chr11:7167064:7167188 |
| *Add3* | NM_013758.4 | SE | 0.08 | chr19:53242504:53242627:chr19:53244310:53244405:chr19:53244998:53247399 |
| *Adrbk1* | NM_130863.2 | RI | -0.06 | chr19:42916414291590:chr19:42913304291239 |
| *Agtpbp1* | NM_023328.3 | MXE | 0.08 | chr13:59532013:59532076:chr13:59531057:59531203:chr13:59528387:59528518:chr13:59525146:59525239 |
| *Agtpbp1* | NM_023328.3 | A5SS | 0.06 | chr13:59557286:59557098\|59557223:chr13:59544387:59544451 |
| *Anks1b* | | SE | 0.05 | chr10:90897247:90897426:chr10:90914696:90914767:chr10:90921296:90921424 |
| *Anks1b* | | A3SS | -0.37 | chr10:90897247:90897426:chr10:90914693\|90914696:90914767 |
| *Arhgap26* | NM_175164.4 | RI | -0.11 | chr18:3935755339357663:chr18:3935779939357828 |
| *Arhgap26* | NM_175164.4 | A5SS | 0.05 | chr18:39357553:39357663\|39357828:chr18:39363117:39363208 |
| *Astn2* | NM_019514.3 | A3SS | 0.1 | chr4:65912561:65912635:chr4:65911785\|65911797:65911660 |
| *Atm* | NM_007499.2 | A5SS | -0.06 | chr9:53451689:53451549\|53451660:chr9:53450518:53450634: |
| *Atp2a3* | NM_016745.3 | SE | 0.13 | chr11:72988970:72989087:chr11:72989475:72989547:chr11:72991689:72993043: |
| *Atp2a3* | NM_016745.3 | SE | 0.09 | chr11:72988970:72989087:chr11:72989475:72989560:chr11:72991689:72993043: |
| *Atp2a3* | NM_016745.3 | A3SS | -0.06 | chr11:72980127:72980345:chr11:72980439\|72980493:72980774: |
| *Atxn2* | | SE | -0.07 | chr5:121802948:121803080:chr5:121806222:121806275:chr5:121810851:121810996: |
| *Auts2* | | MXE | 0.09 | chr5:131445399:131445482:chr5:131443756:131443805:chr5:131443456:131443678:chr5:131437682:131440693: |
| *Auts2* | | A5SS | -0.11 | chr5:131472479:131472238\|131472259:chr5:131470414:131470458: |
| *Bzrap1* | NM_172449.2 | SE | -0.13 | chr11:87781945:87782049:chr11:87782660:87782710:chr11:87784736:87785928: |
| *Bzrap1* | NM_172449.2 | RI | 0.06 | chr11:8778168287781753:chr11:8778194587782049: |
| *Bzrap1* | NM_172449.2 | A5SS | 0.05 | chr11:87781302:87781334\|87781370:chr11:87781490:87781593: |
| *Cacna1a* | NM_007578.3 | MXE | 0.14 | chr8:84612285:84612412:chr8:84614695:84614791:chr8:84615574:84615670:chr8:84617792:84617897: |
| *Cacna1b* | NM_007579.3 | A3SS | -0.14 | chr2:24685768:24685874:chr2:24679640\|24679643:24678876: |
| *Cacna1c* | | MXE | 0.2 | chr6:118675820:118675949:chr6:118674555:118674614:chr6:118674346:118674405:chr6:118670350:118670456: |
| *Cacna1c* | | A5SS | -0.11 | chr6:118630431:118630225\|118630399:chr6:118627442:118627570: |
| *Cacna1d* | | MXE | 0.25 | chr14:30095318:30095428:chr14:30089822:30089905:chr14:30089296:30089379:chr14:30082952:30082996: |
| *Cacna1d* | NM_028981.3 | A3SS | 0.1 | chr14:30130117:30130204:chr14:30128798\|30129848:30128772: |
| *Cacna1g* | NM_009783.3 | A5SS | 0.21 | chr11:94425977:94425764\|94425785:chr11:94418819:94418970: |
| *Cacna1g* | NM_009783.3 | A5SS | 0.11 | chr11:94425977:94425764\|94425785:chr11:94423671:94423724: |
| *Cacna1g* | NM_009783.3 | A3SS | 0.05 | chr11:94416841:94416911:chr11:94416655\|94416704:94416626: |
| *Camk2b* | NM_007595.5 | SE | -0.06 | chr11:5979673:5979721:chr11:5976765:5976893:chr11:5972813:5972888: |
| *Camk2b* | NM_007595.5 | SE | 0.32 | chr11:5988101:5988143:chr11:5987057:5987131:chr11:5982721:5982758: |
| *Camk2b* | NM_007595.5 | RI | 0.09 | chr11:59728885972813:chr11:59727265972632: |
| *Cask* | NM_009806.3 | A5SS | -0.12 | chrX:13786049:13785919\|13785937:chrX:13714666:13714771: |
| *Cask* | NM_009806.3 | A3SS | -0.21 | chrX:13544514:13544629:chrX:13537826\|13537841:13537761: |
| *Ccdc64* | | A5SS | -0.16 | chr5:115652986:115652627\|115652925:chr5:115651819:115651939: |
| *Cdh18* | | A3SS | 0.14 | chr15:23445978:23446095:chr15:23460316\|23460425:23460567: |
| *Chd5* | | SE | -0.19 | chr4:152385433:152385596:chr4:152385724:152385838:chr4:152385949:152386002: |
| *Chd5* | | A3SS | -0.13 | chr4:152384485:152384680:chr4:152385433\|152385534:152385596: |
| *Cntnap5b* | NM_172851.2 | SE | -0.26 | chr1:100369757:100369884:chr1:100370058:100370228:chr1:100379051:100379269: |
| *Cpeb1* | NM_007755.5 | A5SS | 0.13 | chr7:81356998:81356894\|81356909:chr7:81355875:81356011: |
| *Cpeb1* | NM_007755.5 | A3SS | -0.06 | chr7:81436203:81436377:chr7:81372200\|81372203:81372012: |
| *Cpne9* | NM_170673.3 | SE | 0.13 | chr6:113283047:113283087:chr6:113283196:113283242:chr6:113283350:113283453: |
| *Cpne9* | NM_170673.3 | RI | 0.12 | chr6:113292522113293064:chr6:113293709113293755: |
| *Cpne9* | NM_170673.3 | RI | 0.21 | chr6:113292522113292575:chr6:113293003113293064: |
| *Crhr1* | NM_007762.5 | SE | -0.21 | chr11:104173306:104173391:chr11:104173595:104173730:chr11:104173889:104173930: |
| *Crhr1* | NM_007762.5 | SE | -0.13 | chr11:104153521:104153608:chr11:104163804:104163889:chr11:104169056:104169162: |
| *Crhr1* | NM_007762.5 | SE | 0.06 | chr11:104153521:104153608:chr11:104159252:104159371:chr11:104163804:104163889: |
| *Cttnbp2* | | A5SS | 0.13 | chr6:18435446:18433820\|18433983:chr6:18427439:18427642: |
| *Cwf19l1* | | A3SS | -0.15 | chr19:44133000:44133084:chr19:44132148\|44132164:44132086: |
| *Dab1* | NM_010014.3 | RI | 0.13 | chr4:104680070104680108:chr4:104680191104680382: |
| *Ddx26b* | NM_172779.4 | SE | -0.13 | chrX:56493039:56493133:chrX:56494133:56494243:chrX:56496516:56496734: |
| *Ddx26b* | NM_172779.4 | RI | -0.11 | chrX:5649303956493133:chrX:5649651656496734: |
| *Ddx26b* | NM_172779.4 | A5SS | 0.12 | chrX:56506880:56506999\|56507062:chrX:56507214:56507843: |
| *Dgkb* | NM_178681.4 | SE | 0.54 | chr12:38084167:38084243:chr12:38087573:38087593:chr12:38100364:38100517: |
| *Dgkh* | | A5SS | -0.17 | chr14:78604550:78604277\|78604303:chr14:78603026:78603133: |
| *Dgkh* | | A3SS | -0.09 | chr14:78577449:78577557:chr14:78576012\|78576017:78575887: |
| *Dgkz* | NM_138306.2 | RI | 0.06 | chr2:9193408991934056:chr2:9193384291933743: |
| *Dgkz* | NM_138306.2 | A3SS | -0.51 | chr2:91936858:91936937:chr2:91936771\|91936774:91936702: |
| *Dgkz* | NM_138306.2 | A3SS | -0.07 | chr2:91944022:91944099:chr2:91942684\|91942814:91942628: |
| *Dlg2* | | SE | -0.06 | chr7:92417224:92417323:chr7:92420632:92420673:chr7:92427691:92427741: |
| *Dlg2* | | MXE | -0.16 | chr7:92386915:92386990:chr7:92417224:92417323:chr7:92418088:92418133:chr7:92420632:92420673: |
| *Dmxl2* | | A3SS | -0.37 | chr9:54398047:54398123:chr9:54396367\|54396370:54396240: |
| *Ebf1* | NM_007897.3 | A5SS | -0.18 | chr11:44621167:44621230\|44621349:chr11:44621921:44621976: |
| *Ebf1* | NM_007897.3 | A3SS | 0.14 | chr11:44883814:44883931:chr11:44907904\|44907907:44908037: |
| *Ebf2* | NM_010095.6 | A3SS | -0.4 | chr14:67233292:67233956:chr14:67234826\|67234831:67235265: |
| *Efr3a* | | SE | 0.19 | chr15:65787041:65787246:chr15:65788302:65788381:chr15:65790140:65791351: |
| *Egfem1* | | SE | -0.16 | chr3:29657130:29657267:chr3:29662377:29662499:chr3:29668241:29668438: |
| *Egfem1* | | MXE | -0.1 | chr3:29153497:29153601:chr3:29357145:29357195:chr3:29513888:29514010:chr3:29582907:29583029: |
| *Emd* | NM_007927.3 | SE | 0.08 | chrX:74255743:74255876:chrX:74256878:74256924:chrX:74260782:74261548: |

| | | | | |
|---|---|---|---|---|
| *Eml6* | NM_146016.2 | A3SS | 0.3 | chr11:29821639:29821757:chr11:29819092\|29819184:29818906: |
| *Epha5* | NM_007937.3 | A3SS | -0.22 | chr5:84150342:84150501:chr5:84142380\|84142383:84142278: |
| *Epha5* | NM_007937.3 | A3SS | 0.09 | chr5:84331228:84331891:chr5:84237554\|84237556:84237399: |
| *Erc2* | NM_177814.4 | SE | 0.23 | chr14:28302876:28303010:chr14:28317249:28317314:chr14:28475603:28478537: |
| *Erc2* | NM_177814.4 | SE | -0.18 | chr14:27652687:27653483:chr14:27776159:27776182:chr14:27776826:27777242: |
| *Erc2* | NM_177814.4 | SE | 0.09 | chr14:28025033:28025173:chr14:28029434:28029469:chr14:28040343:28040536: |
| *Erc2* | NM_177814.4 | SE | 0.05 | chr14:27622442:27622614:chr14:27623683:27623749:chr14:27652687:27653483: |
| *Etl4* | | SE | -0.14 | chr2:20801542:20801661:chr2:20805585:20805662:chr2:20807902:20808024: |
| *Etl4* | | SE | 0.24 | chr2:20289894:20290041:chr2:20339922:20340109:chr2:20529959:20530242: |
| *Etl4* | | SE | 0.14 | chr2:20798616:20798783:chr2:20801542:20801661:chr2:20807902:20808024: |
| *Etl4* | | SE | -0.12 | chr2:20805585:20805662:chr2:20805775:20807373:chr2:20807902:20808024: |
| *Etl4* | | SE | 0.15 | chr2:20743459:20744291:chr2:20759547:20759651:chr2:20760215:20760264: |
| *Exosc8* | NM_027148.3 | RI | 0.28 | chr3:5473433254734296:chr3:5473416654734103: |
| *Fam149b* | NM_172379.3 | SE | 0.15 | chr14:20356355:20356497:chr14:20358024:20358140:chr14:20363263:20363450: |
| *Fam149b* | NM_172379.3 | SE | 0.08 | chr14:20375510:20375613:chr14:20377775:20377999:chr14:20378400:20378523: |
| *Fmr1* | NM_008013.3 | A3SS | 0.07 | chrX:68710584:68710779:chrX:68712231\|68712306:68712413: |
| *Foxp1* | NM_053202.2 | A3SS | -0.13 | chr6:98945534:98945735:chr6:98945423\|98945426:98945347: |
| *Foxp1* | NM_053202.2 | A3SS | -0.08 | chr6:99016528:99016665:chr6:99015510\|99015513:99015424: |
| *Foxp2* | NM_053242.4 | SE | -0.15 | chr6:14901349:14901761:chr6:14988323:14988464:chr6:15084482:15084575: |
| *Foxp2* | NM_053242.4 | SE | -0.08 | chr6:15405559:15405642:chr6:15409741:15409942:chr6:15411026:15411102: |
| *Foxp2* | NM_053242.4 | A5SS | 0.12 | chr6:15084482:15084575\|15084578:chr6:15196951:15197128: |
| *Foxp2* | NM_053242.4 | A3SS | 0.1 | chr6:15377681:15377730:chr6:15377870\|15377888:15378088: |
| *Foxp2* | NM_053242.4 | A3SS | 0.21 | chr6:15376689:15376826:chr6:15377870\|15377951:15378088: |
| *Foxp2* | NM_053242.4 | A3SS | 0.17 | chr6:15376689:15376826:chr6:15377888\|15377951:15378088: |
| *Frmd4a* | NM_172475.3 | SE | 0.05 | chr2:4605970:4606066:chr2:4607999:4608070:chr2:4610967:4614043: |
| *Frmd5* | NM_172673.3 | A3SS | -0.11 | chr2:121548889:121549232:chr2:121547648\|121548023:121545529: |
| *Gabra1* | NM_010250.5 | RI | -0.06 | chr11:4218293042182874:chr11:4218238942182166: |
| *Garnl3* | | A5SS | 0.06 | chr2:33105023:33104849\|33104853:chr2:33085875:33085944: |
| *Gnptab* | NM_001004164.2 | RI | -0.11 | chr10:8837922488379314:chr10:8837946988379535: |
| *Gpc6* | NM_011821.3 | SE | -0.06 | chr14:117888522:117888665:chr14:117892342:117892371:chr14:117951138:117951274: |
| *Gria3* | NM_016886.4 | SE | 0.06 | chrX:41654006:41654253:chrX:41654810:41654924:chrX:41669501:41669615: |
| *Gria3* | NM_016886.4 | SE | 0.07 | chrX:41654810:41654924:chrX:41669501:41669615:chrX:41672219:41672466: |
| *Gria3* | NM_016886.4 | SE | 0.05 | chrX:41654006:41654253:chrX:41654810:41654924:chrX:41672219:41672466: |
| *Grin1* | NM_008169.3 | A3SS | 0.15 | chr2:25295792:25295937:chr2:25292128\|25292484:25291181: |
| *Grip1* | | SE | 0.24 | chr10:119978438:119978607:chr10:119985474:119986484:chr10:119999752:119999938: |
| *Grip1* | | SE | 0.09 | chr10:119454034:119454422:chr10:119897715:119897795:chr10:119929901:119930036: |
| *Grip1* | NM_028736.2 | A3SS | 0.13 | chr10:119978438:119978607:chr10:119985474\|119986332:119986484: |
| *Gstm1* | NM_010358.5 | A3SS | 0.18 | chr3:108016328:108016428:chr3:108015118\|108015145:108014945: |
| *Hdac9* | NM_024124.3 | A5SS | -0.11 | chr12:34431989:34431863\|34431869:chr12:34429498:34429619: |
| *Hspa14* | NM_015765.2 | A3SS | 0.28 | chr2:3511015:3511097:chr2:3509020\|3509095:3508293: |
| *Htt* | NM_010414.3 | SE | 0.13 | chr5:34782742:34782825:chr5:34790267:34790387:chr5:34794105:34794164: |
| *Huwe1* | | SE | 0.19 | chrX:151857808:151857913:chrX:151859455:151859688:chrX:151860167:151860268: |
| *Huwe1* | | SE | -0.27 | chrX:151803662:151803802:chrX:151804446:151804540:chrX:151806272:151806407: |
| *Huwe1* | | SE | 0.05 | chrX:151803282:151803569:chrX:151803756:151803802:chrX:151806272:151806407: |
| *Huwe1* | | MXE | -0.08 | chrX:151803282:151803569:chrX:151803756:151803802:chrX:151804446:151804540:chrX:151806272:151806407: |
| *Huwe1* | | A5SS | -0.08 | chrX:151923479:151923561\|151923606:chrX:151924662:151924761: |
| *Huwe1* | | A3SS | 0.43 | chrX:151803662:151803802:chrX:151806272\|151806276:151806407: |
| *Inpp4a* | NM_030266.4 | SE | 0.17 | chr1:37379922:37380060:chr1:37383339:37383371:chr1:37387698:37387870: |
| *Inpp4a* | NM_030266.4 | SE | 0.12 | chr1:37372311:37372419:chr1:37374272:37374289:chr1:37377578:37377761: |
| *Inpp4a* | NM_030266.4 | SE | 0.08 | chr1:37372311:37372419:chr1:37374275:37374289:chr1:37377578:37377761: |
| *Inpp5a* | NM_183144.3 | A5SS | -0.17 | chr7:139578182:139578218\|139578269:chr7:139578354:139579652: |
| *Kalrn* | | A5SS | 0.21 | chr16:34256360:34256152\|34256191:chr16:34252225:34252399: |
| *Kalrn* | | A3SS | -0.18 | chr16:34227027:34227146:chr16:34220247\|34220274:34220056: |
| *Kalrn* | | A3SS | 0.06 | chr16:34016392:34016630:chr16:34014211\|34014214:34013958: |
| *Kcnc3* | | SE | -0.09 | chr7:44598384:44598575:chr7:44600869:44600928:chr7:44601869:44604751: |
| *Kcnd3* | NM_019931.1 | SE | 0.05 | chr3:105665457:105665546:chr3:105666962:105667018:chr3:105668080:105668327: |
| *Kcnma1* | NM_010610.3 | A3SS | -0.28 | chr14:23309040:23309264:chr14:23300042\|23300045:23298694: |
| *Kcnma1* | NM_010610.3 | A3SS | -0.15 | chr14:23306725:23306753:chr14:23300042\|23300045:23298694: |
| *Kitl* | NM_013598.2 | SE | 0.12 | chr10:100079974:100080130:chr10:100080857:100080940:chr10:100087347:100087456: |
| *Klc2* | NM_008451.2 | A3SS | -0.12 | chr19:5118122:5118408:chr19:5117432\|5117469:5117194: |
| *Klc2* | NM_008451.2 | A3SS | 0.08 | chr19:5118184:5118298:chr19:5117432\|5117469:5117194: |
| *Lpcat4* | NM_207206.2 | RI | 0.41 | chr2:112242476112242588:chr2:112242701112242761: |
| *Lrp8* | | SE | -0.21 | chr4:107847444:107847566:chr4:107850104:107850142:chr4:107851308:107851427: |
| *Lrp8* | | SE | -0.07 | chr4:107843216:107843344:chr4:107846239:107846619:chr4:107847444:107847566: |
| *Lrrtm4* | | SE | -0.32 | chr6:80018877:80019186:chr6:80019554:80019706:chr6:80021614:80024932: |
| *Lrrtm4* | | A3SS | -0.22 | chr6:80018877:80019186:chr6:80019624\|80019643:80019706: |
| *Macrod2* | | SE | -0.08 | chr2:142210100:142210144:chr2:142217578:142217661:chr2:142256521:142256631: |
| *Macrod2* | | A5SS | 0.2 | chr2:142256521:142256583\|142256631:chr2:142261093:142261161: |
| *Magi2* | NM_015823.3 | SE | 0.13 | chr5:20534337:20534526:chr5:20543585:20543626:chr5:20550207:20550298: |
| *Mdm4* | NM_008575.4 | A3SS | -0.07 | chr1:133018777:133018884:chr1:133012711\|133012714:133012637: |
| *Mef2c* | NM_025282.3 | SE | -0.36 | chr13:83592778:83592981:chr13:83625470:83625607:chr13:83633055:83633241: |
| *Mef2c* | NM_025282.3 | SE | 0.19 | chr13:83652814:83652986:chr13:83654598:83654621:chr13:83655507:83655636: |
| *Mef2c* | NM_025282.3 | MXE | 0.06 | chr13:83592778:83592981:chr13:83625265:83625408:chr13:83625470:83625607:chr13:83633055:83633241: |
| *Mef2c* | | A3SS | 0.08 | chr13:83656217:83656352:chr13:83662307\|83662403:83667079: |
| *Meg3* | | SE | 0.05 | chr12:109546430:109546542:chr12:109549291:109549371:chr12:109549479:109549552: |

| | | | | |
|---|---|---|---|---|
| *Mettl16* | NM_026197.3 | MXE | 0.13 | chr11:74795982:74796124:chr11:74800326:74800455:chr11:74802893:74803012:chr11:74803831:74803900: |
| *Mfsd11* | NM_178620.3 | RI | -0.29 | chr11:116863906116863946:chr11:116865891116865956: |
| *Mfsd11* | NM_178620.3 | A3SS | -0.08 | chr11:116858474:116858581:chr11:116859299\|116859324:116859378: |
| *Mfsd11* | NM_178620.3 | A3SS | 0.16 | chr11:116858474:116858581:chr11:116859324\|116859349:116859378: |
| *Mfsd11* | NM_178620.3 | A3SS | -0.13 | chr11:116858474:116858581:chr11:116859299\|116859349:116859378: |
| *Napb* | NM_019632.3 | RI | 0.11 | chr2:148709433148709317:chr2:148707205148707159: |
| *Napb* | NM_019632.3 | A5SS | 0.21 | chr2:148709433:148707159\|148709317:chr2:148706946:148707023: |
| *Nell1* | | SE | -0.06 | chr7:50701156:50701251:chr7:50826260:50826400:chr7:50848483:50848676: |
| *Nktr* | NM_010918.2 | SE | 0.06 | chr9:121731478:121731565:chr9:121738388:121740075:chr9:121741122:121741151: |
| *Nlgn1* | NM_138666.3 | A5SS | 0.2 | chr3:25440077:25439865\|25439892:chr3:25435853:25436642: |
| *Nrcam* | NM_176930.4 | SE | 0.11 | chr12:44566288:44566412:chr12:44566840:44566869:chr12:44568544:44568645: |
| *Nrcam* | NM_176930.4 | SE | -0.06 | chr12:44532458:44532659:chr12:44535057:44535074:chr12:44537251:44537356: |
| *Nrcam* | NM_176930.4 | SE | 0.15 | chr12:44578145:44578180:chr12:44584835:44584987:chr12:44590108:44590239: |
| *Nrp2* | NM_010939.3 | SE | -0.14 | chr1:62786281:62786316:chr1:62812534:62812584:chr1:62815333:62818692: |
| *Nrp2* | NM_010939.2 | A5SS | 0.06 | chr1:62786281:62786301\|62786316:chr1:62795709:62797091: |
| *Nrp2* | NM_010939.3 | A5SS | 0.19 | chr1:62786281:62786301\|62786316:chr1:62815333:62818692: |
| *Nrxn3* | NM_172544.3 | SE | 0.14 | chr12:89348246:89348449:chr12:89354453:89354479:chr12:89503018:89503137: |
| *Nrxn3* | | SE | 0.06 | chr12:88953390:88953616:chr12:88954811:88954911:chr12:89187022:89187323: |
| *Nrxn3* | | A5SS | 0.06 | chr12:90283572:90283650\|90283659:chr12:90332109:90334933: |
| *Nrxn3* | NM_172544.3 | A5SS | 0.39 | chr12:90283572:90283650\|90283659:chr12:90321936:90322641: |
| *Nrxn3* | | A3SS | -0.24 | chr12:90283572:90283650:chr12:90331788\|90332109:90334933: |
| *P2rx4* | NM_011026.3 | SE | 0.09 | chr5:122727198:122727291:chr5:122727399:122727464:chr5:122727745:122727840: |
| *P2rx4* | NM_011026.3 | SE | -0.07 | chr5:122719130:122719226:chr5:122724594:122724674:chr5:122724990:122725131: |
| *Pbrm1* | | SE | -0.18 | chr14:31054158:31054255:chr14:31056236:31056280:chr14:31061469:31061745: |
| *Pbrm1* | | SE | -0.07 | chr14:31022720:31022859:chr14:31023067:31023170:chr14:31025498:31025650: |
| *Pbrm1* | | SE | 0.05 | chr14:31107109:31107310:chr14:31110415:31110579:chr14:31113849:31114004: |
| *Pbrm1* | | SE | -0.06 | chr14:31019138:31019293:chr14:31022720:31022859:chr14:31023067:31023179: |
| *Pbrm1* | | SE | -0.06 | chr14:31044624:31044709:chr14:31045330:31045425:chr14:31046332:31046423: |
| *Pbrm1* | | A5SS | 0.06 | chr14:31023067:31023170\|31023179:chr14:31025498:31025650: |
| *Pbrm1* | | A3SS | 0.05 | chr14:31074771:31074982:chr14:31082550\|31082553:31082735: |
| *Pcdh19* | XM_205287.3 | A3SS | -0.25 | chrX:133632895:133632953:chrX:133625438\|133625441:133625269: |
| *Pcp2* | | RI | -0.09 | chr8:36246323624508:chr8:36235583623377: |
| *Pcsk6* | | SE | 0.36 | chr7:66025228:66025407:chr7:66025566:66025604:chr7:66031749:66031851: |
| *Pde9a* | NM_008804.4 | SE | 0.09 | chr17:31415280:31415357:chr17:31420245:31420288:chr17:31443166:31443220: |
| *Pde9a* | NM_008804.4 | SE | -0.07 | chr17:31420245:31420288:chr17:31443166:31443220:chr17:31443838:31443908: |
| *Pde9a* | NM_008804.4 | RI | -0.09 | chr17:3145990131460005:chr17:3146017931460261: |
| *Phactr2* | | MXE | -0.09 | chr10:13261766:13261924:chr10:13257577:13257816:chr10:13255310:13255321:chr10:13253342:13253879: |
| *Pkp4* | NM_026361.2 | SE | -0.1 | chr2:59347924:59348041:chr2:59350494:59350622:chr2:59352262:59352335: |
| *Pkp4* | | SE | 0.06 | chr2:59214678:59214814:chr2:59266388:59266500:chr2:59289218:59289252: |
| *Pkp4* | | MXE | 0.08 | chr2:59214678:59214814:chr2:59266388:59266500:chr2:59289218:59289252:chr2:59305066:59305197: |
| *Pkp4* | NM_026361.2 | A3SS | -0.06 | chr2:59305305:59305492:chr2:59308008\|59308085:59308557: |
| *Pkp4* | NM_026361.2 | A3SS | 0.06 | chr2:59310102:59310290:chr2:59311694\|59311697:59311913: |
| *Plcb1* | NM_019677.2 | SE | -0.07 | chr2:135387798:135387884:chr2:135399249:135399366:chr2:135472053:135475258: |
| *Polrmt* | NM_172551.3 | RI | -0.11 | chr10:7973750979737457:chr10:7973736279737264: |
| *Ppp1r1b* | NM_144828.2 | RI | -0.11 | chr11:9834999698350110:chr11:9835056998350629: |
| *Ppp4r4* | NM_028980.3 | SE | -0.1 | chr12:103603972:103604058:chr12:103604577:103604692:chr12:103604967:103605060: |
| *Ppp4r4* | NM_028980.3 | SE | -0.07 | chr12:103558368:103558470:chr12:103575709:103575781:chr12:103576281:103576428: |
| *Prkcg* | NM_011102.4 | SE | -0.14 | chr7:3314512:3314541:chr7:3318860:3319012:chr7:3319531:3319719: |
| *Ptbp1* | NM_008956.3 | A3SS | 0.05 | chr10:79854605:79854721:chr10:79856339\|79856504:79856534: |
| *Rabggtb* | NM_011231.2 | A3SS | -0.12 | chr3:153909349:153909459:chr3:153908899\|153908915:153908790: |
| *Rasal2* | | A3SS | 0.11 | chr1:157161125:157161326:chr1:157158837\|157158858:157158659: |
| *Rbfox2* | NM_053104.6 | A3SS | 0.06 | chr15:77099240:77099332:chr15:77098072\|77098084:77097940: |
| *Rbms3* | NM_178660.4 | MXE | 0.17 | chr9:116678647:116678697:chr9:116636369:116636479:chr9:116586018:116586095:chr9:116572747:116578702: |
| *Rell2* | | SE | 0.15 | chr18:37956929:37956994:chr18:37957589:37957774:chr18:37958014:37958389: |
| *Rell2* | | SE | 0.08 | chr18:37955559:37955909:chr18:37956328:37956567:chr18:37956929:37956994: |
| *Rell2* | | A5SS | 0.15 | chr18:37955139:37955916\|37956567:chr18:37956929:37956994: |
| *Rell2* | | A3SS | -0.37 | chr18:37956929:37956994:chr18:37957459\|37957589:37957774: |
| *Reln* | NM_011261.2 | A3SS | 0.08 | chr5:21895989:21896087:chr5:21891567\|21891697:21891562: |
| *Rgs8* | NM_026380.3 | SE | -0.11 | chr1:153665836:153665916:chr1:153667825:153667953:chr1:153670783:153670884: |
| *Rims2* | | SE | -0.08 | chr15:39511264:39511374:chr15:39517806:39517871:chr15:39534838:39535017: |
| *Rims2* | | SE | 0.1 | chr15:39585590:39585762:chr15:39610064:39610141:chr15:39616269:39616510: |
| *Rims2* | NM_053271.2 | MXE | -0.06 | chr15:39436997:39437922:chr15:39452040:39452107:chr15:39452224:39452432:chr15:39454360:39454479: |
| *Scn1a* | | A5SS | -0.07 | chr2:66324951:66324571\|66324655:chr2:66323312:66323444: |
| *Shank1* | | SE | 0.41 | chr7:44333543:44333618:chr7:44334033:44334059:chr7:44342098:44342180: |
| *Shank1* | | SE | -0.15 | chr7:44343658:44343738:chr7:44344298:44344321:chr7:44344506:44344574: |
| *Shank1* | | A3SS | -0.27 | chr7:44342098:44342180:chr7:44342420\|44342425:44342544: |
| *Slc12a5* | NM_020333.2 | SE | 0.12 | chr2:164996322:164996518:chr2:164996864:164996878:chr2:164997110:164997243: |
| *Slc6a8* | NM_133987.2 | A5SS | -0.15 | chrX:73679919:73680047\|73680071:chrX:73680132:73680234: |
| *Slc9a1* | NM_016981.2 | A3SS | -0.29 | chr4:133419454:133419543:chr4:133419925\|133419947:133419995: |
| *Snap25* | NM_011428.3 | MXE | -0.08 | chr2:136763573:136763621:chr2:136769743:136769860:chr2:136770057:136770174:chr2:136773895:136774020: |
| *Snhg11* | NM_175692.3 | SE | 0.65 | chr2:158376081:158376326:chr2:158376745:158376824:chr2:158376994:158377072: |
| *Sox5* | NM_011444.3 | A3SS | -0.13 | chr6:144023169:144023237:chr6:143960917\|143960920:143960797: |
| *Spock3* | NM_023689.3 | SE | -0.23 | chr8:62951800:62951988:chr8:63113434:63113442:chr8:63113543:63113588: |
| *Stam* | NM_011484.2 | SE | -0.06 | chr2:14074112:14074336:chr2:14102401:14102485:chr2:14115817:14115892: |
| *Stat3* | XM_011248846.2 | A5SS | 0.05 | chr11:100893286:100893074\|100893077:chr11:100889886:100889928: |

| | | | | |
|---|---|---|---|---|
| *Sugp2* | | A5SS | -0.06 | chr8:70260469:70260589\|70261317:chr8:70262871:70263105: |
| *Svil* | NM_153153.3 | SE | 0.1 | chr18:5059230:5059369:chr18:5060515:5060610:chr18:5062162:5062405: |
| *Svil* | | SE | 0.28 | chr18:5037108:5037165:chr18:5040148:5040239:chr18:5046589:5046646: |
| *Svil* | NM_153153.3 | SE | 0.26 | chr18:5055980:5056849:chr18:5057275:5057439:chr18:5058125:5058159: |
| *Svil* | | SE | -0.06 | chr18:4920540:4920874:chr18:4971090:4971197:chr18:5037108:5037165: |
| *Syn1* | NM_013680.4 | A3SS | 0.19 | chrX:20862499:20863090:chrX:20861571\|20861609:20860511: |
| *Tbp* | NM_013684.3 | SE | 0.1 | chr17:15513530:15513621:chr17:15514230:15514397:chr17:15515616:15515710: |
| *Tbp* | NM_013684.3 | SE | -0.3 | chr17:15499888:15500024:chr17:15501154:15501202:chr17:15502962:15503128: |
| *Tbp* | NM_013684.3 | SE | -0.08 | chr17:15499888:15500024:chr17:15501154:15501202:chr17:15502965:15503128: |
| *Tbp* | NM_013684.3 | SE | -0.17 | chr17:15499888:15500024:chr17:15501154:15501202:chr17:15501312:15501357: |
| *Tbp* | NM_013684.3 | SE | 0.07 | chr17:15501154:15501202:chr17:15501312:15501357:chr17:15502965:15503128: |
| *Tbp* | NM_013684.3 | MXE | -0.09 | chr17:15499888:15500024:chr17:15501154:15501202:chr17:15501312:15501357:chr17:15502965:15503128: |
| *Tbp* | NM_013684.3 | A5SS | 0.08 | chr17:15501154:15501202\|15501357:chr17:15502965:15503128: |
| *Tbp* | NM_013684.3 | A3SS | 0.06 | chr17:15501154:15501202:chr17:15502962\|15502965:15503128: |
| *Tenm2* | | A5SS | -0.22 | chr11:36139761:36139549\|36139576:chr11:36106719:36106865: |
| *Tenm2* | | A3SS | 0.05 | chr11:36141483:36141683:chr11:36139758\|36139761:36139576: |
| *Tenm4* | | SE | 0.6 | chr7:96553478:96553747:chr7:96643999:96644208:chr7:96694770:96695025: |
| *Tenm4* | NM_011858.4 | SE | 0.28 | chr7:96852333:96852582:chr7:96854717:96854737:chr7:96863450:96863682: |
| *Tenm4* | | SE | -0.17 | chr7:96436760:96436858:chr7:96496244:96496340:chr7:96550012:96550299: |
| *Tenm4* | NM_011858.4 | SE | 0.14 | chr7:96694770:96695025:chr7:96702642:96702740:chr7:96703926:96704161: |
| *Trim9* | | A5SS | -0.1 | chr12:70251260:70250957\|70251011:chr12:70248265:70248427: |
| *Ttll5* | NM_00181423.2 | SE | -0.12 | chr12:86020521:86020603:chr12:86024220:86024342:chr12:86053143:86053760: |
| *Ttll5* | | SE | 0.06 | chr12:86012648:86012862:chr12:86014767:86014859:chr12:86020521:86020603: |
| *Ttll5* | | A3SS | -0.08 | chr12:85933282:85933674:chr12:85939326\|85939329:85939510: |
| *Unc5d* | NM_153135.3 | A5SS | -0.09 | chr8:28683252:28683103\|28683163:chr8:28675275:28675439: |
| *Unc5d* | NM_153135.3 | A5SS | -0.27 | chr8:28724406:28724221\|28724260:chr8:28719617:28719994: |
| *Vps13d* | | A3SS | 0.08 | chr4:145178189:145178379:chr4:145178077\|145178095:145178012: |
| *Zfand4* | | SE | -0.12 | chr6:116273573:116273872:chr6:116284740:116284815:chr6:116285854:116285921: |

SE:skipping exon

RI: retention intron

MXE: mutually exclusive exons

A5SS: Alternative 5’ splice sites

A3SS: Alternative 3’ splice sites

**Table S3. DEGs in PC by scRNA-seq analysis (mutant/control)**

| Gene | P_value | Avg_log2FC | PC in mutant mouse | PC in control mouse |
|---|---|---|---|---|
| | | | Population of expressing cells | Population of expressing cells |
| *Pvalb* | 1.51E-06 | -1.41 | 0.45 | 0.80 |
| *Meg3* | 2.19E-16 | -1.06 | 1.00 | 1.00 |
| *Snhg11* | 1.53E-14 | -0.92 | 0.99 | 1.00 |
| *Fxyd7* | 1.50E-10 | -0.86 | 0.67 | 0.98 |
| *Scg5* | 2.21E-15 | -0.82 | 0.69 | 0.98 |
| *Prkcg* | 3.40E-07 | -0.82 | 0.87 | 0.96 |
| *Epha4* | 0.000310 | -0.81 | 0.75 | 0.91 |
| *Epha7* | 0.000816 | -0.78 | 0.55 | 0.78 |
| *Chgb* | 7.83E-11 | -0.75 | 0.87 | 1.00 |
| *C1qtnf4* | 1.72E-10 | -0.73 | 0.96 | 1.00 |
| *Cacna2d2* | 1.28E-09 | -0.72 | 0.99 | 1.00 |
| *Map1a* | 2.06E-10 | -0.72 | 0.87 | 1.00 |
| *Atp2b2* | 8.12E-09 | -0.71 | 1.00 | 1.00 |
| *Lars2* | 5.65E-10 | -0.71 | 0.66 | 0.95 |
| *Gad2* | 5.86E-07 | -0.71 | 0.72 | 0.93 |
| *Kcnc3* | 1.10E-06 | -0.71 | 0.97 | 1.00 |
| *Oxct1* | 5.70E-12 | -0.70 | 1.00 | 1.00 |
| *Nav1* | 1.59E-13 | -0.68 | 1.00 | 1.00 |
| *Car8* | 9.27E-15 | -0.66 | 1.00 | 1.00 |
| *Washc2* | 1.54E-05 | -0.64 | 0.84 | 0.95 |
| *Kcnma1* | 1.05E-06 | -0.64 | 0.97 | 1.00 |
| *Eif2s3y* | 3.96E-11 | -0.64 | 0.31 | 0.84 |
| *Shank1* | 1.25E-10 | -0.64 | 0.99 | 1.00 |
| *Dst* | 5.21E-07 | -0.63 | 0.93 | 1.00 |
| *Malat1* | 0.000270 | -0.63 | 1.00 | 1.00 |
| *Synpr* | 3.16E-05 | -0.63 | 0.39 | 0.67 |
| *Lpgat1* | 5.78E-06 | -0.62 | 0.96 | 1.00 |
| *Sema7a* | 3.30E-07 | -0.61 | 0.58 | 0.95 |
| *Plxna4* | 6.27E-09 | -0.60 | 0.55 | 0.91 |
| *Grm1* | 7.37E-05 | -0.59 | 0.97 | 0.98 |
| *Kifc2* | 2.59E-10 | -0.59 | 0.61 | 0.91 |
| *Adcy1* | 1.32E-07 | -0.58 | 1.00 | 1.00 |
| *Atp1b1* | 1.13E-06 | -0.58 | 1.00 | 1.00 |
| *Nexn* | 1.02E-08 | -0.58 | 0.93 | 0.98 |
| *Rgs7bp* | 3.52E-08 | -0.56 | 0.97 | 1.00 |
| *Cntnap5a* | 0.00581 | -0.56 | 0.82 | 0.86 |
| *Ktn1* | 8.37E-09 | -0.56 | 0.93 | 1.00 |
| *Ptprr* | 6.32E-06 | -0.56 | 0.82 | 1.00 |
| *Inpp5a* | 1.24E-06 | -0.55 | 0.99 | 1.00 |

| | | | | |
|---|---|---|---|---|
| *Cttnbp2* | 7.51E-07 | -0.55 | 0.87 | 0.96 |
| *Ccdc88a* | 6.68E-11 | -0.55 | 1.00 | 1.00 |
| *Cacna1g* | 1.58E-06 | -0.55 | 0.81 | 1.00 |
| *Syne1* | 7.24E-08 | -0.54 | 0.76 | 0.98 |
| *Camk2a* | 8.90E-07 | -0.54 | 0.72 | 0.95 |
| *AI593442* | 0.000864 | -0.53 | 0.99 | 1.00 |
| *Calr* | 8.54E-07 | -0.52 | 0.97 | 1.00 |
| *Atrnl1* | 8.90E-08 | -0.52 | 0.67 | 0.95 |
| *Npnt* | 0.00201 | -0.52 | 0.33 | 0.55 |
| *Mia3* | 6.70E-06 | -0.52 | 0.85 | 0.95 |
| *Cd47* | 2.04E-05 | -0.52 | 0.99 | 1.00 |
| *Ndnf* | 0.00806 | -0.52 | 0.87 | 0.95 |
| *Hist1h2ap* | 0.0151 | -0.52 | 0.30 | 0.58 |
| *Atp2a2* | 4.16E-06 | -0.51 | 1.00 | 1.00 |
| *Tpr* | 1.22E-07 | -0.51 | 0.96 | 1.00 |
| *Arhgef33* | 5.72E-09 | -0.51 | 0.99 | 1.00 |
| *Kcna1* | 1.79E-06 | -0.51 | 0.67 | 0.87 |
| *Thy1* | 5.58E-06 | -0.51 | 0.97 | 1.00 |
| *Itpr1* | 0.000551 | -0.51 | 1.00 | 1.00 |
| *Rps27a* | 1.55E-05 | 0.50 | 1.00 | 1.00 |
| *Sf3b5* | 4.18E-06 | 0.51 | 0.96 | 0.95 |
| *Polr2k* | 2.15E-06 | 0.51 | 0.93 | 0.82 |
| *Nop10* | 6.65E-06 | 0.52 | 0.99 | 0.96 |
| *Cck* | 0.0218 | 0.52 | 0.84 | 0.75 |
| *Rpl24* | 1.71E-06 | 0.54 | 0.99 | 1.00 |
| *Ccng1* | 1.16E-05 | 0.54 | 0.69 | 0.46 |
| *Rbp1* | 0.00289 | 0.54 | 0.66 | 0.42 |
| *Rps3a1* | 1.24E-06 | 0.55 | 1.00 | 1.00 |
| *Nnat* | 0.0949 | 0.55 | 0.73 | 0.76 |
| *Tbca* | 8.67E-09 | 0.55 | 1.00 | 0.98 |
| *Bcl2l11* | 0.000314 | 0.55 | 0.78 | 0.60 |
| *Rpl3* | 3.15E-07 | 0.56 | 1.00 | 1.00 |
| *Scg2* | 0.00360 | 0.56 | 0.79 | 0.69 |
| *Tmsb4x* | 5.09E-05 | 0.56 | 1.00 | 1.00 |
| *Rps26* | 1.21E-07 | 0.56 | 1.00 | 1.00 |
| *Rps28* | 1.03E-07 | 0.56 | 1.00 | 1.00 |
| *Atp6v1g1* | 6.58E-07 | 0.57 | 0.97 | 1.00 |
| *Gm32913* | 0.0534 | 0.57 | 0.12 | 0.02 |
| *Jund* | 2.62E-05 | 0.57 | 1.00 | 0.98 |
| *Rpl37a* | 6.73E-08 | 0.58 | 1.00 | 1.00 |
| *Lsm7* | 3.89E-10 | 0.58 | 0.99 | 0.95 |
| *Rps27l* | 1.70E-06 | 0.59 | 0.96 | 0.95 |

| | | | | |
|---|---|---|---|---|
| *Rps4x* | 1.20E-05 | 0.59 | 0.99 | 1.00 |
| *Rpl35* | 3.96E-07 | 0.60 | 0.99 | 1.00 |
| *Zbtb20* | 0.00374 | 0.60 | 0.63 | 0.55 |
| *Rpl18a* | 8.62E-12 | 0.60 | 1.00 | 1.00 |
| *Banf1* | 4.57E-10 | 0.60 | 1.00 | 0.95 |
| *Rpl39* | 3.40E-10 | 0.61 | 1.00 | 1.00 |
| *Marcks* | 6.70E-05 | 0.61 | 0.99 | 0.98 |
| *Serf1* | 9.31E-08 | 0.61 | 0.97 | 0.98 |
| *Rpl38* | 3.98E-10 | 0.63 | 1.00 | 1.00 |
| *Ptgds* | 0.624 | 0.63 | 0.39 | 0.46 |
| *Dbpht2* | 8.02E-05 | 0.63 | 0.91 | 0.82 |
| *Rps27* | 1.78E-09 | 0.63 | 1.00 | 1.00 |
| *Btg1* | 6.36E-07 | 0.64 | 1.00 | 0.96 |
| *Rps15a* | 2.47E-11 | 0.64 | 1.00 | 1.00 |
| *Mtpn* | 7.90E-08 | 0.64 | 0.97 | 1.00 |
| *Rps29* | 1.68E-09 | 0.64 | 1.00 | 1.00 |
| *Slc5a7* | 6.43E-06 | 0.65 | 0.46 | 0.07 |
| *Rps5* | 7.90E-10 | 0.66 | 1.00 | 1.00 |
| *Taf1d* | 1.08E-07 | 0.66 | 0.99 | 0.96 |
| *Snrpd3* | 2.40E-09 | 0.69 | 0.96 | 0.98 |
| *Rpl37* | 3.41E-11 | 0.71 | 1.00 | 1.00 |
| *Gm10076* | 3.19E-10 | 0.71 | 0.97 | 0.96 |
| *Sec61g* | 2.43E-12 | 0.72 | 1.00 | 1.00 |
| *Nsa2* | 6.75E-11 | 0.74 | 0.99 | 0.96 |
| *1500015O10Rik* | 0.00615 | 0.78 | 0.45 | 0.20 |
| *Bax* | 9.35E-12 | 0.80 | 0.99 | 0.93 |
| *Sat1* | 4.74E-08 | 0.80 | 0.87 | 0.82 |
| *Cirbp* | 1.13E-14 | 0.82 | 1.00 | 1.00 |
| *Cdkn1a* | 2.03E-05 | 0.89 | 0.63 | 0.33 |

**Table S4. DEGs in PC by snRNA-seq analysis (mutant/control)**

| Gene | P_value | Avg_log2FC | PC in mutant mouse | PC in control mouse |
|---|---|---|---|---|
| | | | Population of expressing cells | Population of expressing cells |
| *Paxbp1* | 6.78E-12 | -1.11 | 0.87 | 1.00 |
| *Hnrnpa2b1* | 2.92E-13 | -0.91 | 0.86 | 0.99 |
| *Gm26906* | 8.37E-13 | -0.89 | 0.34 | 0.84 |
| *Dcun1d5* | 9.97E-12 | -0.86 | 0.41 | 0.88 |
| *Cntnap2* | 0.00503 | -0.82 | 0.62 | 0.81 |
| *Gm28928* | 0.0311 | -0.81 | 0.16 | 0.29 |
| *Fras1* | 2.30E-05 | -0.79 | 0.66 | 0.88 |
| *Acss2* | 1.47E-10 | -0.79 | 0.30 | 0.75 |
| *Gm40841* | 2.20E-10 | -0.78 | 0.49 | 0.94 |
| *A230006K03Rik* | 5.34E-05 | -0.78 | 0.37 | 0.64 |
| *Epha7* | 0.000906 | -0.78 | 0.52 | 0.78 |
| *Srsf2* | 1.62E-10 | -0.73 | 0.41 | 0.90 |
| *Gm44813* | 9.19E-09 | -0.67 | 0.18 | 0.61 |
| *Kcnip1* | 5.81E-05 | -0.66 | 0.86 | 0.99 |
| *Taco1* | 8.16E-08 | -0.65 | 0.11 | 0.54 |
| *Gm26917* | 0.290 | -0.62 | 0.51 | 0.69 |
| *Tra2a* | 1.35E-07 | -0.61 | 0.66 | 0.95 |
| *Lrrtm4* | 0.00130 | -0.60 | 0.80 | 0.94 |
| *Aff1* | 1.79E-06 | -0.60 | 0.34 | 0.74 |
| *mt-Nd4l* | 0.00741 | -0.58 | 0.24 | 0.49 |
| *Myef2* | 3.88E-07 | -0.58 | 0.61 | 0.91 |
| *Hnrnpdl* | 3.63E-07 | -0.58 | 0.49 | 0.86 |
| *Hs6st3* | 0.00570 | -0.57 | 0.78 | 0.93 |
| *Sos1* | 1.34E-05 | -0.57 | 0.86 | 0.98 |
| *Apip* | 2.93E-09 | -0.57 | 0.21 | 0.68 |
| *Adarb2* | 0.593 | -0.57 | 0.10 | 0.12 |
| *Gria2* | 3.17E-10 | -0.56 | 1.00 | 1.00 |
| *Dpp10* | 3.17E-05 | -0.56 | 1.00 | 0.99 |
| *Zfp711* | 1.41E-06 | -0.56 | 0.34 | 0.71 |
| *Phldb2* | 5.40E-05 | -0.56 | 0.24 | 0.59 |
| *Camk1d* | 1.19E-06 | -0.55 | 0.62 | 0.92 |
| *Setbp1* | 4.54E-05 | -0.55 | 0.73 | 0.94 |
| *1110019D14Rik* | 0.000308 | -0.54 | 0.38 | 0.69 |
| *9630028H03Rik* | 0.000615 | -0.54 | 0.87 | 0.97 |
| *Cdh13* | 3.46E-05 | -0.54 | 0.51 | 0.84 |
| *Epha4* | 0.00240 | -0.54 | 0.48 | 0.71 |
| *Ttc14* | 6.69E-07 | -0.54 | 0.70 | 0.97 |
| *Ppib* | 3.72E-07 | -0.54 | 0.30 | 0.74 |
| *Polb* | 6.65E-06 | -0.54 | 0.80 | 0.98 |

| | | | | |
|---|---|---|---|---|
| *Kcna1* | 7.00E-06 | -0.53 | 0.44 | 0.76 |
| *Akap7* | 4.37E-06 | -0.53 | 0.37 | 0.75 |
| *4732471J01Rik* | 3.46E-06 | -0.53 | 0.48 | 0.85 |
| *Lhfpl3* | 0.000620 | -0.53 | 0.82 | 0.94 |
| *Csmd1* | 0.326 | -0.53 | 0.54 | 0.57 |
| *Ddx3y* | 1.84E-11 | -0.53 | 0.00 | 0.43 |
| *Large1* | 3.36E-05 | -0.52 | 0.93 | 1.00 |
| *Olfm3* | 2.82E-06 | -0.52 | 0.04 | 0.29 |
| *Rbm25* | 3.91E-07 | -0.52 | 0.80 | 0.99 |
| *Cnot10* | 1.21E-06 | -0.52 | 0.32 | 0.76 |
| *Cdc42bpa* | 4.46E-08 | -0.52 | 0.97 | 1.00 |
| *Pak1* | 0.000253 | -0.52 | 0.39 | 0.71 |
| *Prpf39* | 1.31E-07 | -0.52 | 0.48 | 0.94 |
| *Zfp57* | 1.60E-05 | -0.52 | 0.37 | 0.71 |
| *Inpp5a* | 5.89E-05 | -0.51 | 0.90 | 0.99 |
| *Htr2c* | 0.0100 | -0.51 | 0.14 | 0.31 |
| *Arpp21* | 0.000334 | -0.51 | 0.32 | 0.69 |
| *Atr* | 8.29E-06 | -0.51 | 0.31 | 0.71 |
| *Rab10* | 3.59E-05 | -0.50 | 0.52 | 0.85 |
| *Nrxn2* | 0.00162 | 0.50 | 0.78 | 0.76 |
| *Zmiz2* | 0.000383 | 0.50 | 0.42 | 0.26 |
| *Rpl41* | 0.0258 | 0.50 | 0.63 | 0.60 |
| *Ascc2* | 0.00189 | 0.50 | 0.56 | 0.50 |
| *Dcc* | 0.494 | 0.50 | 0.41 | 0.54 |
| *Ccdc107* | 0.0107 | 0.51 | 0.72 | 0.75 |
| *Runx1t1* | 0.0334 | 0.51 | 0.38 | 0.36 |
| *Dcx* | 0.000516 | 0.51 | 0.80 | 0.71 |
| *1110051M20Rik* | 0.000748 | 0.51 | 0.82 | 0.79 |
| *Arhgdig* | 0.00112 | 0.51 | 0.47 | 0.33 |
| *Lrrc28* | 0.000675 | 0.51 | 0.47 | 0.32 |
| *Mcf2l* | 0.00162 | 0.51 | 0.75 | 0.78 |
| *Edc4* | 0.000834 | 0.51 | 0.45 | 0.31 |
| *Dynlt1a* | 0.000381 | 0.51 | 0.55 | 0.38 |
| *Nell1os* | 0.0670 | 0.51 | 0.32 | 0.28 |
| *Gm10658* | 0.000699 | 0.52 | 0.37 | 0.21 |
| *Zfp523* | 0.000180 | 0.52 | 0.54 | 0.35 |
| *Cop1* | 0.000534 | 0.52 | 0.69 | 0.62 |
| *Sfmbt2* | 0.00259 | 0.52 | 0.56 | 0.53 |
| *Tspoap1* | 0.000109 | 0.52 | 0.83 | 0.68 |
| *P2ry14* | 0.0197 | 0.53 | 0.69 | 0.66 |
| *Epha8* | 0.00256 | 0.53 | 0.47 | 0.37 |
| *Snx32* | 0.00604 | 0.53 | 0.80 | 0.81 |

| | | | | |
|---|---|---|---|---|
| *Gm12353* | 0.000826 | 0.53 | 0.99 | 0.97 |
| *Clpp* | 0.00387 | 0.53 | 0.44 | 0.37 |
| *Cul9* | 0.00147 | 0.53 | 0.62 | 0.54 |
| *Fam216a* | 0.00197 | 0.53 | 0.48 | 0.36 |
| *Gm48383* | 0.00370 | 0.53 | 0.65 | 0.58 |
| *Rps27* | 0.0319 | 0.53 | 0.65 | 0.69 |
| *Comtd1* | 0.000707 | 0.54 | 0.51 | 0.39 |
| *2610020C07Rik* | 0.00446 | 0.54 | 0.66 | 0.63 |
| *Nsun5* | 0.00306 | 0.54 | 0.45 | 0.35 |
| *Flii* | 0.000365 | 0.54 | 0.63 | 0.42 |
| *Tmem145* | 0.000625 | 0.54 | 0.78 | 0.74 |
| *Nisch* | 0.00127 | 0.54 | 0.92 | 0.95 |
| *Maged1* | 0.000561 | 0.54 | 0.56 | 0.42 |
| *Sympk* | 0.00266 | 0.54 | 0.62 | 0.55 |
| *Lrp8* | 0.000641 | 0.54 | 0.68 | 0.60 |
| *Nkain2* | 0.613 | 0.54 | 0.35 | 0.43 |
| *Gm26907* | 0.000423 | 0.54 | 0.30 | 0.10 |
| *Idh3g* | 0.000154 | 0.54 | 0.44 | 0.23 |
| *Phf12* | 0.00250 | 0.55 | 0.66 | 0.65 |
| *mt-Cytb* | 0.0131 | 0.55 | 0.92 | 0.88 |
| *Coro1b* | 0.000796 | 0.55 | 0.58 | 0.49 |
| *Drg1* | 0.00142 | 0.55 | 0.83 | 0.85 |
| *Lrrk1* | 0.000268 | 0.55 | 0.25 | 0.04 |
| *Gm20388* | 3.76E-07 | 0.55 | 1.00 | 0.99 |
| *Asap2* | 0.00106 | 0.55 | 0.73 | 0.69 |
| *Dlgap2* | 0.00188 | 0.55 | 0.80 | 0.79 |
| *Agrn* | 2.19E-05 | 0.56 | 0.69 | 0.51 |
| *Cobl* | 0.134 | 0.56 | 0.23 | 0.18 |
| *mt-Nd3* | 0.00463 | 0.56 | 0.58 | 0.46 |
| *Neurl4* | 0.00669 | 0.56 | 0.49 | 0.53 |
| *Chd5* | 0.000572 | 0.56 | 0.66 | 0.57 |
| *Tm9sf4* | 0.000839 | 0.56 | 0.72 | 0.63 |
| *Rev1* | 0.00108 | 0.56 | 0.75 | 0.69 |
| *Gm16183* | 6.85E-05 | 0.56 | 0.94 | 0.90 |
| *Plxna3* | 6.00E-05 | 0.56 | 0.44 | 0.21 |
| *Uqcrc1* | 0.000455 | 0.56 | 0.65 | 0.57 |
| *Dtx3* | 0.00115 | 0.56 | 0.72 | 0.75 |
| *Gm26733* | 2.17E-05 | 0.56 | 0.92 | 0.85 |
| *Lypla2* | 1.95E-06 | 0.56 | 0.48 | 0.14 |
| *Dcaf15* | 0.00293 | 0.56 | 0.51 | 0.40 |
| *Gm48742* | 0.00324 | 0.57 | 0.55 | 0.45 |
| *Kcnt2* | 0.225 | 0.57 | 0.41 | 0.46 |

| | | | | |
|---|---|---|---|---|
| *Leng8* | 0.00197 | 0.58 | 0.79 | 0.83 |
| *Grip1os2* | 0.00277 | 0.58 | 0.69 | 0.62 |
| *Gm45645* | 0.00166 | 0.58 | 0.52 | 0.36 |
| *Gnptg* | 0.000600 | 0.58 | 0.66 | 0.60 |
| *Usp36* | 9.61E-05 | 0.58 | 0.49 | 0.30 |
| *Lrrc45* | 0.000622 | 0.58 | 0.54 | 0.42 |
| *Cacna2d2* | 4.82E-05 | 0.59 | 0.99 | 0.98 |
| *Abca2* | 4.96E-05 | 0.59 | 0.51 | 0.29 |
| *Cadm1* | 0.0529 | 0.59 | 0.56 | 0.58 |
| *Rps6kb2* | 0.000676 | 0.59 | 0.76 | 0.72 |
| *Slc22a17* | 3.91E-06 | 0.60 | 0.75 | 0.54 |
| *Klc2* | 0.000167 | 0.60 | 0.61 | 0.46 |
| *Rap1gap* | 7.85E-05 | 0.60 | 0.65 | 0.54 |
| *Celf3* | 0.000412 | 0.60 | 0.65 | 0.57 |
| *Col4a1* | 0.000487 | 0.60 | 0.66 | 0.50 |
| *Klhdc10* | 7.89E-05 | 0.60 | 0.89 | 0.88 |
| *Atxn2l* | 3.82E-05 | 0.60 | 0.83 | 0.80 |
| *Tenm2* | 5.02E-05 | 0.61 | 0.97 | 0.92 |
| *Gtf2ird1* | 0.000693 | 0.61 | 0.92 | 0.92 |
| *Parp1* | 0.00109 | 0.61 | 0.61 | 0.58 |
| *Strn4* | 0.000298 | 0.61 | 0.70 | 0.61 |
| *Stx1a* | 7.37E-05 | 0.61 | 0.62 | 0.43 |
| *Kcnb2* | 4.08E-05 | 0.61 | 0.99 | 0.98 |
| *Erbb4* | 0.136 | 0.62 | 0.30 | 0.29 |
| *Dlc1* | 0.0372 | 0.62 | 0.18 | 0.14 |
| *Tiam1* | 0.00507 | 0.62 | 0.63 | 0.61 |
| *Brd2* | 1.03E-05 | 0.63 | 0.45 | 0.19 |
| *Gm26904* | 0.000103 | 0.63 | 0.87 | 0.91 |
| *1600010M07Rik* | 0.0283 | 0.63 | 0.66 | 0.66 |
| *Snrpf* | 1.02E-06 | 0.63 | 0.73 | 0.52 |
| *Egfem1* | 0.0140 | 0.63 | 0.72 | 0.59 |
| *Apba3* | 5.97E-05 | 0.64 | 0.41 | 0.19 |
| *Ttyh1* | 0.000764 | 0.64 | 0.61 | 0.60 |
| *Atp5b* | 2.09E-05 | 0.64 | 0.76 | 0.69 |
| *Adamtsl1* | 0.0968 | 0.64 | 0.54 | 0.47 |
| *Leng9* | 2.88E-05 | 0.65 | 0.54 | 0.35 |
| *Gm26518* | 0.000156 | 0.65 | 0.78 | 0.72 |
| *Rundc3a* | 1.72E-05 | 0.65 | 0.61 | 0.40 |
| *Sag* | 7.23E-05 | 0.65 | 0.62 | 0.47 |
| *Gm26749* | 1.22E-06 | 0.65 | 0.83 | 0.71 |
| *Ggt7* | 4.81E-05 | 0.66 | 0.75 | 0.68 |
| *Itga11* | 3.76E-05 | 0.67 | 0.54 | 0.31 |

| | | | | |
|---|---|---|---|---|
| *Rnpepl1* | 7.21E-06 | 0.67 | 0.61 | 0.41 |
| *Pycard* | 5.68E-07 | 0.68 | 0.48 | 0.14 |
| *Mapk8ip1* | 0.000191 | 0.68 | 0.69 | 0.60 |
| *mt-Co1* | 0.00948 | 0.68 | 0.90 | 0.91 |
| *Ssr4* | 3.43E-05 | 0.68 | 0.65 | 0.52 |
| *Gm42439* | 1.45E-06 | 0.68 | 1.00 | 0.97 |
| *Gm49179* | 8.03E-06 | 0.69 | 0.83 | 0.70 |
| *Mpped2* | 0.0389 | 0.69 | 0.42 | 0.34 |
| *Car10* | 0.165 | 0.69 | 0.37 | 0.39 |
| *Gpc6* | 0.0528 | 0.70 | 0.62 | 0.55 |
| *Gm28376* | 0.000681 | 0.70 | 0.85 | 0.79 |
| *Syt7* | 9.48E-06 | 0.70 | 0.76 | 0.61 |
| *Tecr* | 2.92E-08 | 0.70 | 0.99 | 0.95 |
| *Gm14211* | 0.0292 | 0.71 | 0.70 | 0.83 |
| *Tmem208* | 1.28E-05 | 0.71 | 0.68 | 0.60 |
| *Thoc1* | 5.70E-10 | 0.71 | 0.96 | 0.91 |
| *Arglu1* | 1.53E-08 | 0.73 | 1.00 | 0.98 |
| *mt-Nd1* | 0.00111 | 0.73 | 0.86 | 0.74 |
| *C130071C03Rik* | 4.94E-05 | 0.74 | 0.93 | 0.90 |
| *Basp1* | 5.77E-07 | 0.75 | 0.90 | 0.88 |
| *D830024N08Rik* | 2.38E-05 | 0.75 | 0.73 | 0.64 |
| *Gm35188* | 5.65E-06 | 0.75 | 1.00 | 0.98 |
| *Ssbp4* | 1.91E-06 | 0.75 | 0.66 | 0.41 |
| *4930445B16Rik* | 0.00208 | 0.75 | 0.79 | 0.74 |
| *Gripap1* | 2.67E-07 | 0.75 | 0.75 | 0.56 |
| *Ptprs* | 3.35E-08 | 0.76 | 0.99 | 0.98 |
| *Ttll4* | 2.96E-07 | 0.77 | 0.75 | 0.58 |
| *Fndc9* | 1.02E-05 | 0.77 | 0.76 | 0.61 |
| *Ptprn* | 5.68E-09 | 0.77 | 0.97 | 0.91 |
| *Chkb* | 2.80E-05 | 0.78 | 0.51 | 0.34 |
| *Epn1* | 7.02E-05 | 0.79 | 0.52 | 0.36 |
| *Sycp2* | 8.53E-07 | 0.79 | 0.70 | 0.49 |
| *Ckb* | 0.000267 | 0.80 | 0.90 | 0.93 |
| *Exoc4* | 0.000174 | 0.80 | 0.87 | 0.97 |
| *Slc6a1* | 3.47E-07 | 0.80 | 0.92 | 0.85 |
| *Dalrd3* | 7.97E-11 | 0.82 | 1.00 | 0.95 |
| *Clstn2* | 0.233 | 0.82 | 0.42 | 0.51 |
| *Socs6* | 1.15E-07 | 0.82 | 0.59 | 0.30 |
| *Col4a2* | 0.000639 | 0.83 | 0.56 | 0.45 |
| *mt-Co2* | 0.000599 | 0.83 | 0.89 | 0.83 |
| *Celf5* | 6.99E-07 | 0.84 | 0.72 | 0.51 |
| *Gm28375* | 2.00E-06 | 0.84 | 0.72 | 0.57 |

| | | | | |
|---|---|---|---|---|
| *Plekhn1* | 4.16E-09 | 0.86 | 0.79 | 0.54 |
| *Ddx5* | 9.59E-08 | 0.86 | 0.92 | 0.86 |
| *Dvl3* | 2.18E-07 | 0.88 | 0.85 | 0.75 |
| *Gm16105* | 8.22E-10 | 0.90 | 0.58 | 0.16 |
| *Fabp7* | 0.000441 | 0.91 | 0.58 | 0.42 |
| *Sulf1* | 0.00134 | 0.92 | 0.30 | 0.17 |
| *Ppp2r3a* | 7.76E-09 | 0.95 | 0.92 | 0.83 |
| *Cars2* | 9.78E-09 | 0.96 | 0.68 | 0.43 |
| *Zbtb20* | 0.000442 | 0.98 | 0.48 | 0.36 |
| *Nrbp2* | 4.03E-08 | 0.99 | 0.75 | 0.60 |
| *mt-Atp6* | 6.45E-05 | 1.00 | 0.93 | 0.89 |
| *Cpt1c* | 1.73E-10 | 1.08 | 0.90 | 0.78 |
| *Npdc1* | 4.79E-15 | 1.18 | 0.86 | 0.49 |
| *Grk2* | 2.11E-11 | 1.24 | 0.63 | 0.17 |
| *E4f1* | 7.73E-12 | 1.28 | 0.76 | 0.50 |
| *Ano3* | 8.24E-07 | 1.32 | 0.55 | 0.32 |
| *Acsl6* | 3.43E-15 | 1.33 | 0.87 | 0.53 |
| *Kirrel3* | 0.0223 | 1.40 | 0.38 | 0.35 |

**Table S5. DEGs in GC/GCP by scRNA-seq analysis (mutant/control)**

| Gene | P_value | avg_log2FC | GC/GCP in mutant mouse | GC/GCP in control mouse |
|---|---|---|---|---|
| | | | Population of expressing cells | Population of expressing cells |
| *Hist1h2ap* | 2.36E-193 | -1.15 | 0.46 | 0.68 |
| *Hist1h4d* | 8.36E-189 | -0.80 | 0.39 | 0.55 |
| *Mki67* | 1.77E-205 | -0.79 | 0.49 | 0.68 |
| *Hist1h1b* | 9.24E-121 | -0.73 | 0.48 | 0.66 |
| *Smc4* | 6.11E-236 | -0.71 | 0.50 | 0.68 |
| *Nasp* | 0 | -0.71 | 0.80 | 0.92 |
| *Hist1h1e* | 1.12E-175 | -0.67 | 0.62 | 0.76 |
| *Gm42418* | 0 | -0.62 | 1.00 | 1.00 |
| *Ubc* | 0 | -0.60 | 0.69 | 0.86 |
| *Rpl36al* | 0 | -0.60 | 0.70 | 0.88 |
| *Cenpe* | 2.07E-143 | -0.58 | 0.29 | 0.44 |
| *Cenpf* | 2.10E-101 | -0.58 | 0.45 | 0.61 |
| *Sfrp1* | 2.88E-273 | -0.58 | 0.64 | 0.83 |
| *Npm1* | 0 | -0.55 | 0.94 | 0.98 |
| *Rps6* | 0 | -0.55 | 0.92 | 0.97 |
| *Tuba1b* | 4.44E-224 | -0.54 | 0.88 | 0.94 |
| *Prc1* | 3.12E-104 | -0.54 | 0.32 | 0.46 |
| *Hist1h3c* | 4.56E-158 | -0.53 | 0.21 | 0.35 |
| *Lmnb1* | 1.24E-262 | -0.53 | 0.82 | 0.92 |
| *Hist1h2an* | 5.02E-315 | -0.53 | 0.13 | 0.33 |
| *Tpr* | 0 | -0.53 | 0.75 | 0.90 |
| *Ccnd1* | 4.25E-160 | -0.53 | 0.47 | 0.66 |
| *Mcm3* | 7.72E-212 | -0.51 | 0.35 | 0.54 |
| *Cct5* | 0 | -0.51 | 0.73 | 0.89 |
| *1810022K09Rik* | 0 | -0.51 | 0.57 | 0.78 |
| *Smc1a* | 2.57E-322 | -0.51 | 0.70 | 0.87 |
| *Neurod2* | 1.34E-92 | 0.50 | 0.55 | 0.45 |
| *Ptprs* | 3.33E-235 | 0.50 | 0.97 | 0.93 |
| *Camk2n1* | 3.40E-110 | 0.51 | 0.28 | 0.15 |
| *Apoe* | 4.36E-238 | 0.51 | 0.57 | 0.34 |
| *Fabp7* | 1.46E-236 | 0.51 | 0.52 | 0.30 |
| *Jph4* | 4.83E-116 | 0.51 | 0.26 | 0.12 |
| *Ppp3ca* | 2.32E-131 | 0.51 | 0.93 | 0.90 |
| *Pcp4* | 8.28E-48 | 0.52 | 0.22 | 0.16 |
| *Gpm6a* | 2.08E-171 | 0.52 | 0.92 | 0.88 |
| *Rufy3* | 5.81E-179 | 0.53 | 0.89 | 0.82 |
| *Snhg9* | 2.40E-271 | 0.53 | 0.60 | 0.38 |
| *Anks1b* | 2.04E-127 | 0.54 | 0.35 | 0.20 |
| *Serf1* | 5.19E-289 | 0.54 | 0.92 | 0.84 |

| | | | | |
|---|---|---|---|---|
| *Dpysl2* | 6.91E-298 | 0.54 | 0.96 | 0.90 |
| *Prkcb* | 8.20E-118 | 0.55 | 0.45 | 0.31 |
| *Zic1* | 1.88E-256 | 0.55 | 1.00 | 1.00 |
| *Cnr1* | 3.44E-65 | 0.56 | 0.28 | 0.21 |
| *Hist3h2ba* | 8.02E-173 | 0.57 | 0.73 | 0.59 |
| *Dner* | 1.13E-143 | 0.58 | 0.38 | 0.22 |
| *Rtn1* | 8.80E-157 | 0.58 | 0.99 | 0.99 |
| *Gm26735* | 3.97E-217 | 0.59 | 0.46 | 0.25 |
| *Syt1* | 2.31E-87 | 0.59 | 0.44 | 0.35 |
| *mt-Nd4l* | 0 | 0.60 | 0.99 | 0.99 |
| *Zbtb18* | 2.56E-198 | 0.60 | 0.92 | 0.86 |
| *Tmsb4x* | 1.33E-273 | 0.61 | 1.00 | 1.00 |
| *Snca* | 3.01E-119 | 0.61 | 0.41 | 0.28 |
| *Cbln1* | 4.13E-163 | 0.61 | 0.46 | 0.28 |
| *Slc24a5* | 0 | 0.62 | 0.69 | 0.47 |
| *Thra* | 2.55E-166 | 0.62 | 0.63 | 0.46 |
| *Gng3* | 1.50E-151 | 0.62 | 0.63 | 0.49 |
| *Celf4* | 4.37E-119 | 0.63 | 0.57 | 0.44 |
| *Btg1* | 0 | 0.64 | 0.92 | 0.82 |
| *Nrxn3* | 1.43E-184 | 0.68 | 0.36 | 0.16 |
| *Camk2d* | 2.60E-165 | 0.70 | 0.56 | 0.40 |
| *Ly6h* | 1.42E-139 | 0.70 | 0.26 | 0.12 |
| *Nnat* | 3.34E-53 | 0.71 | 0.87 | 0.86 |
| *Meg3* | 1.78E-112 | 0.73 | 0.38 | 0.21 |
| *Cirbp* | 0 | 0.74 | 0.96 | 0.87 |
| *Nrep* | 3.49E-252 | 0.74 | 0.93 | 0.88 |
| *mt-Atp8* | 0 | 0.76 | 0.99 | 0.97 |
| *Tsix* | 0 | 0.76 | 0.33 | 0.00 |
| *Mapt* | 4.36E-166 | 0.79 | 0.42 | 0.25 |
| *Gm10076* | 0 | 0.79 | 0.89 | 0.82 |
| *Dcx* | 1.01E-317 | 0.81 | 0.91 | 0.83 |
| *Mndal* | 0 | 0.81 | 0.38 | 0.00 |
| Gm2694 | 0 | 0.93 | 0.87 | 0.69 |

Table S6. List of primers, ss-donor oligonucleotides and LNA probe

| Nucleotides | Manufacture | Sequence |
|---|---|---|
| hSNUPN_EcoRI_Fw | Eurofins Genomics | GAATTC GAAGAGTTGAGTCAGGCCCTG |
| hSNUPN_BamHI_Rv | Eurofins Genomics | GATGCCTCATGGAGAATTAA GGATCC |
| R55W crRNA | FASMAC | UUUCUCUUCCAGCAAGCGGU |
| R55W ss-oligo | Eurofins Genomics | TCCCCACTCTCCATACCTGTCCAGTCATCTTCAGCCAGTCT<br>TCTGGCATGGTTAACATAATCTAACCACTTCCTGGAAGAG<br>AAAAAGATAGGATCAGGTCTATACAT |
| I312Y* crRNA | FASMAC | GCUCAAUAAUCUGUUGGAGC |
| I312Y* ss-oligo | Eurofins Genomics | GCCCACTGACCACCAAGCCAGAATACGCTGGCCCAGCACC<br>AACAGATTTATTGAGCACAAGAGGATCCAGGAAGACACAA<br>AGGAGAAACTCACACACA |
| R55W Fw | Eurofins Genomics | TGCAGAAATCCAAGTGGCTGGATTATGT |
| R55W Rv | Eurofins Genomics | ACATAATCCAGCCACTTGGATTTCTGCA |
| I312Y* Fw | Eurofins Genomics | TTATGGAGCACAAGAAGAGC |
| I312Y* Rv | Eurofins Genomics | ATCTGCTGGAGCTGGTG |
| LNA probe for<br>U1 SnRNA in ISH | WAKENYAKU | GG(L)TAT(L)CT5(L)CC5(L)TGC5(L)CAG(L)GTA(L)AGT(L)AT |
| Cacna2d2 Fw | Eurofins Genomics | GGGTGCCTATAAGGAGTACCAACTAC |
| Cacna2d2 Rv | Eurofins Genomics | CTTCAGGAACTCGGTGTTGTTGTC |
| Camk2b Fw | Eurofins Genomics | GTTACTCCTGAAGCCAAAAACCTC |
| Camk2b Rv | Eurofins Genomics | CCCGTCCACTGGGTTATGGATAAC |
| Ccdc85a Fw | Eurofins Genomics | CAACCTTGTCCTATGTTAGGCAGC |
| Ccdc85a Rv | Eurofins Genomics | GAGCAATGTACAATCACTCCATGTG |
| Inpp4a Fw | Eurofins Genomics | GAGAAAGTGTGGCTGAATGTGGAC |
| Inpp4a Rv | Eurofins Genomics | GATACTGCAGGCTGAGGTACGAG |
| Lrg8 Fw | Eurofins Genomics | GAGAATGAGTTCCAGTGTGGGGAT |
| Lrg8 Rv | Eurofins Genomics | GGTCAGTGTATCCATCTCATAGCC |
| Prkcg Fw | Eurofins Genomics | ACAAGTTACTGAACCAGGAGGAGG |
| Prkcg Rv | Eurofins Genomics | TCTCTACGAGGGTGCAGTCTACAT |
| Rbms3 Fw | Eurofins Genomics | ATGCAGCCAACTAACATCGTGG |
| Rbms3 Rv | Eurofins Genomics | TTGGACTGTTGGAAGGAGTATGCC |
| Rictor Fw | Eurofins Genomics | AGCTACACAAGCTCTAGAGATGCC |
| Rictor Rv | Eurofins Genomics | GAACCTCCATATGTAGCACTGGAC |
| Slc12a5 Fw | Eurofins Genomics | ACCTGGACCAAGGATAAGTCAGTG |
| Slc12a5 Rv | Eurofins Genomics | TATTCACGATGACCTCGTTCAGCC |
| Snhg11 Fw | Eurofins Genomics | CTCTTGACACCAAGGATGACGAC |
| Snhg11 Rv | Eurofins Genomics | ACTGATGTTGGGGAGTACGTTCCT |
| Ttll5 Fw | Eurofins Genomics | TTGGCAGCCAGACACTACCTAACT |
| Ttll5 Rv | Eurofins Genomics | CCTTGGAGTCTCTGGTTCATCTGT |

**Table S7. List of primary antibodies used in the study**

| **Antibodies** | **Source** | **Identifier** |
|---|---|---|
| Anti-m3G-Cap/m7G-cap | Merck Millipore | Cat# MABE419 |
| Anti-Calbindin D28K | Santa Cruz | Cat# sc-7691 |
| Anti-Calbindin D28K | Frontier Institute | Cat# MSFR100400 |
| Alexa fluor 647® Anti-Coilin [1H10] | abcam | Cat# ab196714 |
| Anti-CRMP2 | ECM biosciences | Cat# CP2161 |
| Anti-CRMP2-pT555 | ECM biosciences | Cat# CP5391 |
| Anti-NeuroD1 | abcam | Cat# ab213725 |
| Anti-Pax2 | abcam | Cat# ab79389 |
| Anti-PKCγ | Frontier Institute | Cat# MSFR104840 |
| Anti-RNUT1 (Snurportin-1) | Abnova | Cat# H00010073-B01 |
| Anti-SMN | Novus | Cat# NB100-1936SS |
| Anti-SNRPD2 | RayBiotech | Cat# 144-06983-50 |
| Anti-vGlut2 | abcam | Cat# ab79157 |
| Anti-Ki67 | Thermo (invitrogen) | Cat# 14-5698-82 |
| Anti-Laminin a2 | ALEXIS | Cat# 804-190-C100 |
| Anti-GFP | Clontech | Cat# 632592 |

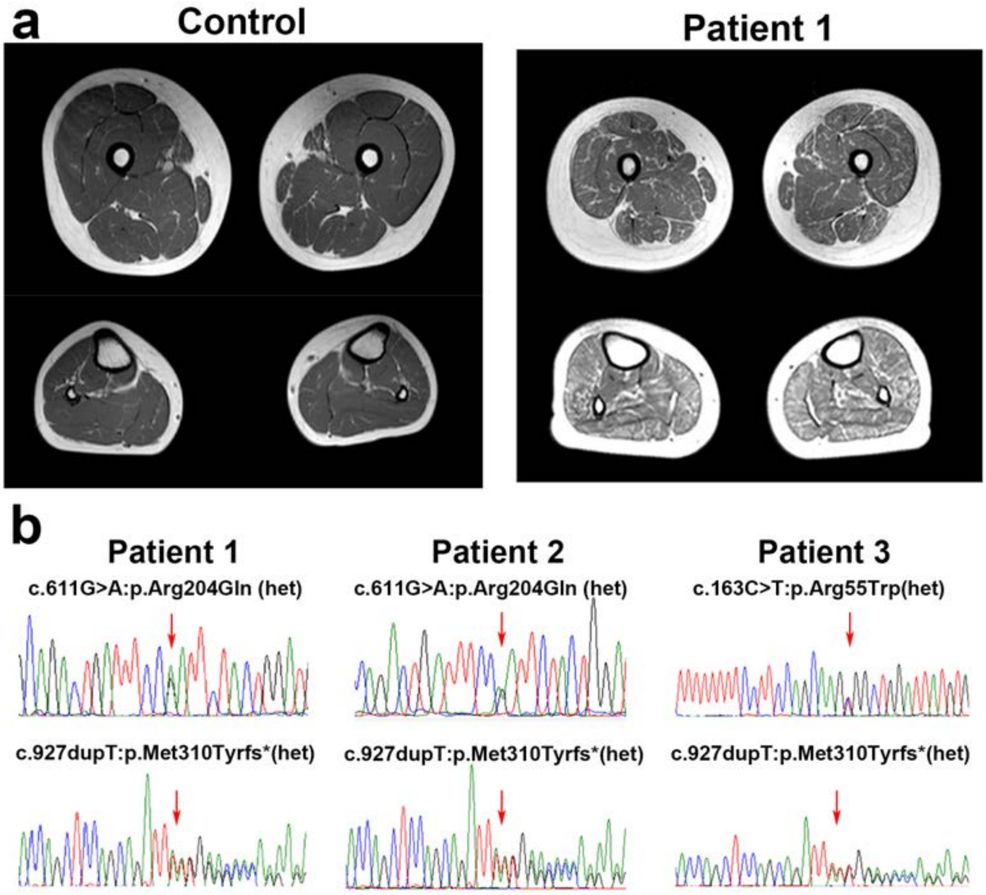


**Supplementary Figure 1. Muscle image and *SNUPN* gene sequence for patients.**

(a) T1W muscle imaging of control and Patient 1 at the age range of 6-10 years-old. For Patient 1, diffuse fatty infiltration is seen in thigh and leg muscles. (b) Compound heterozygous c.611G>A and c.927dupT were identified in Patient 1 and 2, and c.163C>T and c.927dupT in Patient 3 in *SNUPN* gene.

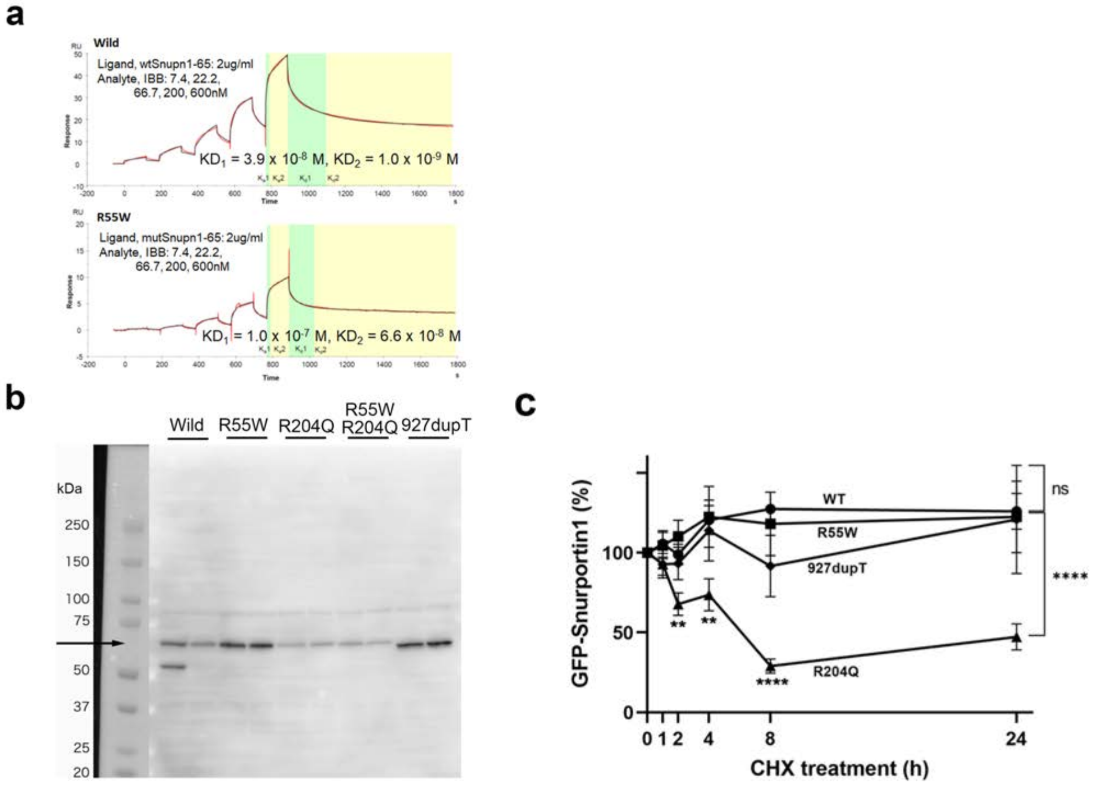


**Supplementary Figure 2. Binding of N-terminal peptides of R55W snurportin-1 to importin beta and the stability of EGFP-snurportin-1 with/without mutations**

(a) Binding of recombinant N-terminal peptides of snurportin-1 and importin beta on BIACORE. Upper, wild type; Lower, R55W. (b) Expression of various EGFP-snurportin-1 proteins in HeLa cells at 24 hours after transfection with CHX treatment (c) Pulse-chase stabilities with CHX treatment of various EGFP-snurportin-1 proteins in HeLa cells after transfection.

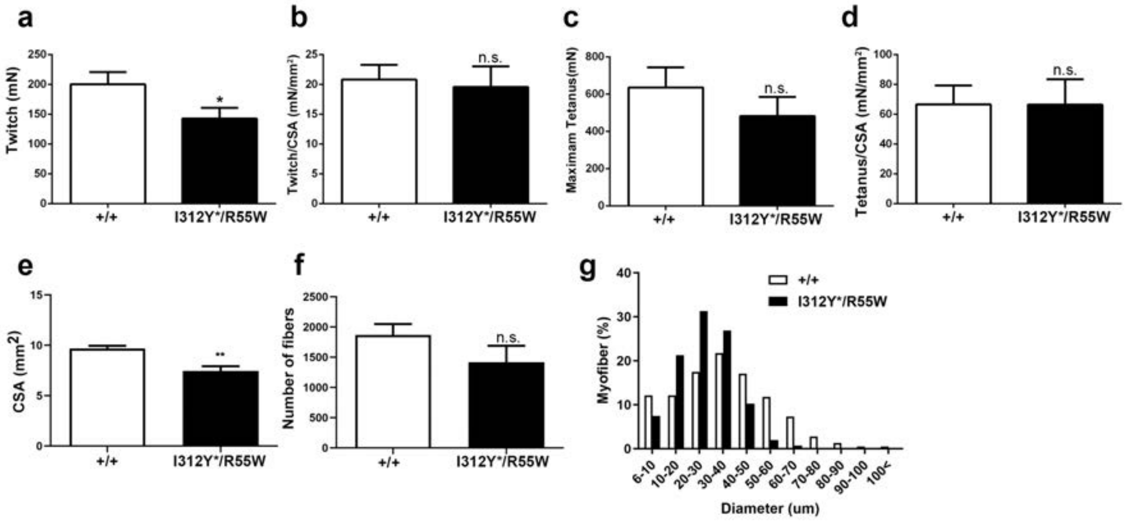


**Supplementary Figure 3. Muscle force and pathology of I312Y*/R55W**

(a, b) Contractile properties of the gastrocnemius muscle, as determined by *ex vivo* measurement of specific isometric force (mN). It was corrected by Cross-Sectional Area (CSA: $mm^2$), and specific tetanic force (c, d). (e) The area of tibialis anterior (TA) muscle for wild type and I312Y*/R55W mice. **$p<0.01$ (f) The total number of muscle fibers. (g) The rate of fiber size for wild type and I312Y*/R55W. (N=3 for each genotype)

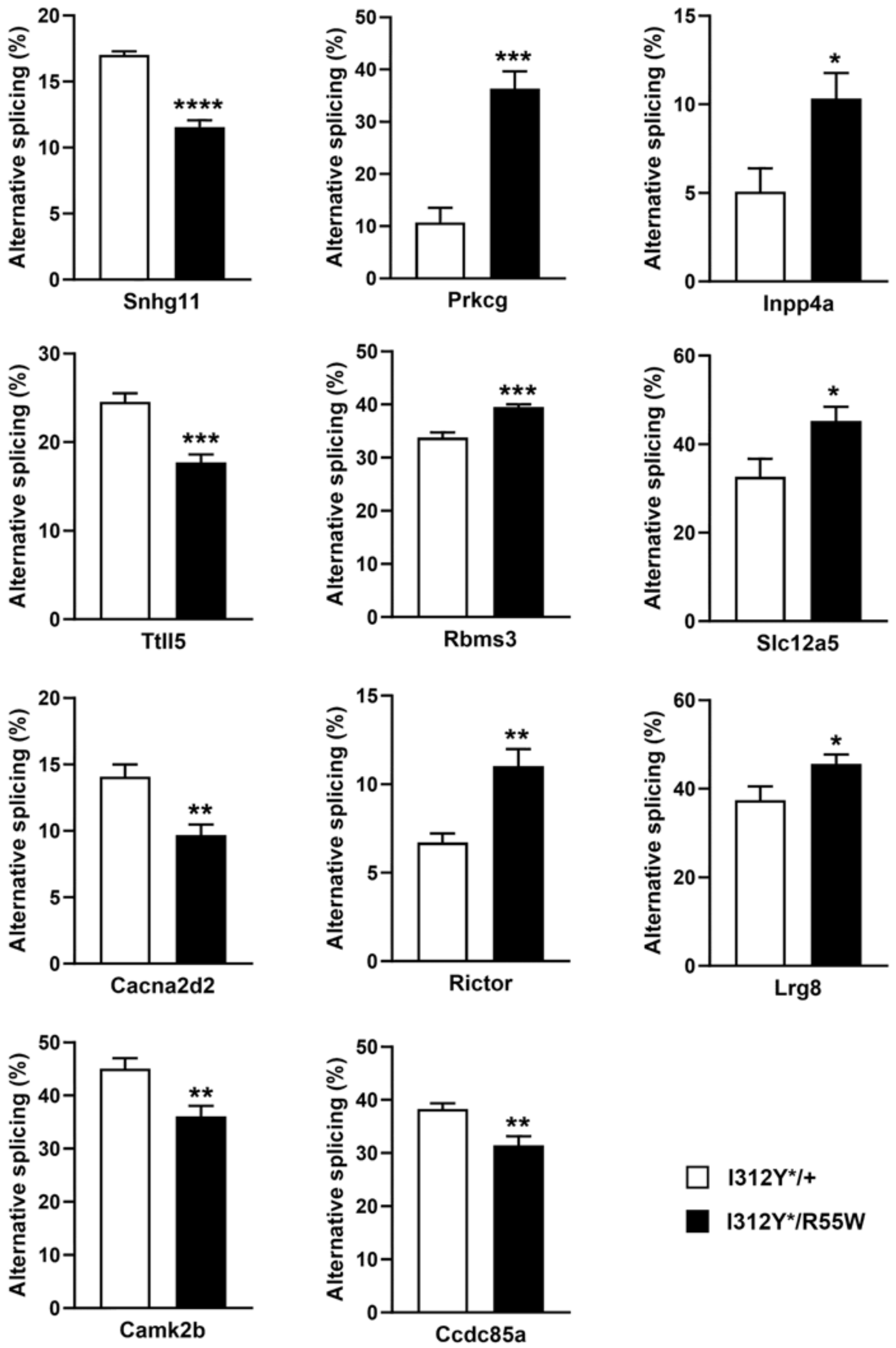


**Supplementary Figure 4. Splicing alteration of the transcripts of 11 PC-specific genes by RT-PCR**

Alternative splicing ratio of each gene between I312Y*/+ and I312Y*/R55W was shown.

*$p < 0.05$, **$p < 0.01$, ***$p < 0.001$, ****$p < 0.0001$

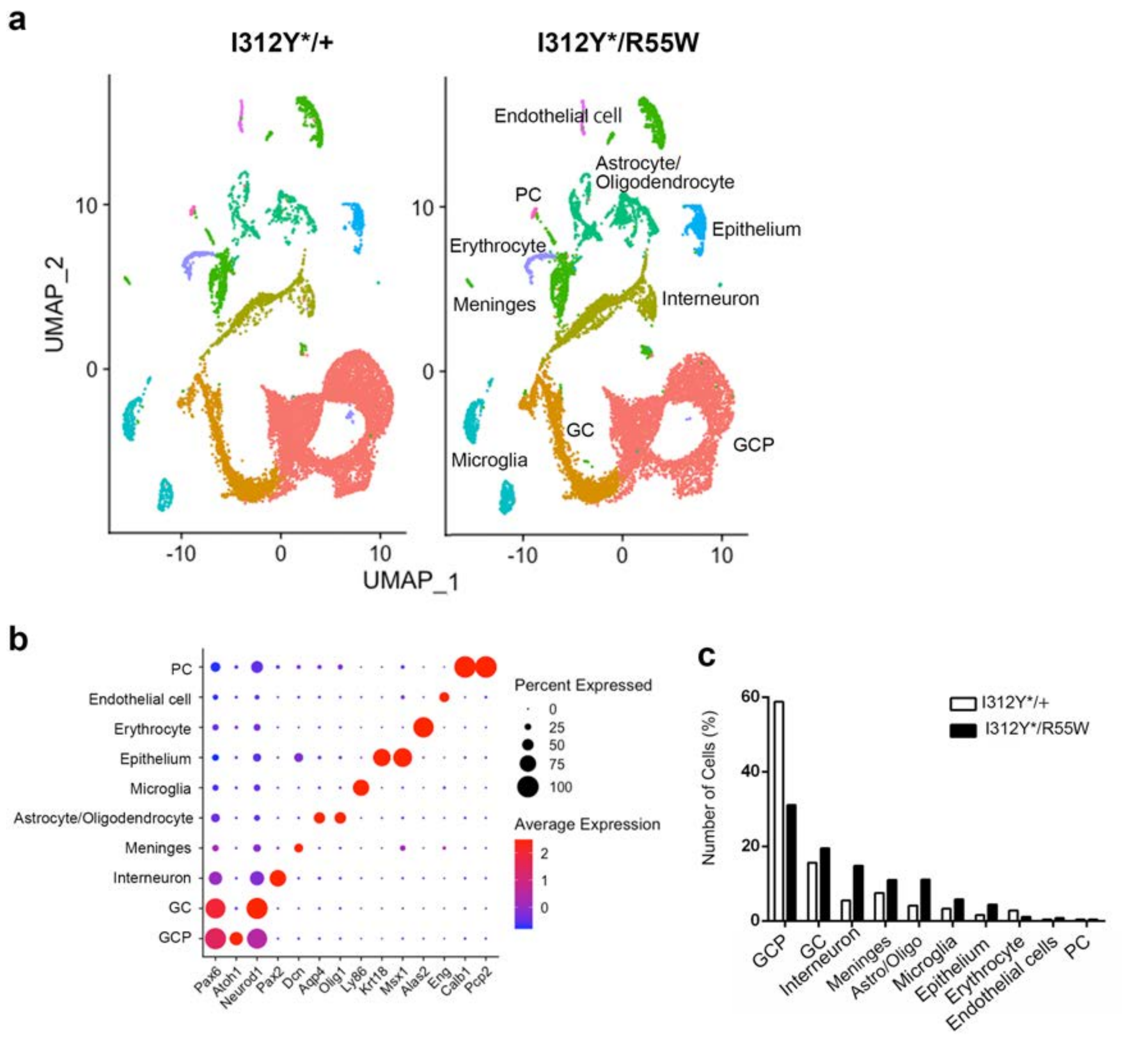


**Supplementary Figure 5. scRNA-seq analysis for whole cerebellum of I312Y*/+ and I312Y*/R55W**

(a) Two-dimensional UMAP distribution of cell clusters of both genotypes. Note the large overlap between control (I312*Y/+) and mutant (I312Y*/R55W). UMAP divided into 10 clustered by R-analysis (resolution=0.05). (b) Identification of cell clusters based on the expression of known genes. (c) Percentage of cell types in the cerebellum of P6 control and mutant mice.

GCP: Granular Cell Progenitor, GC: Granular cell, PC: Purkinje Cell

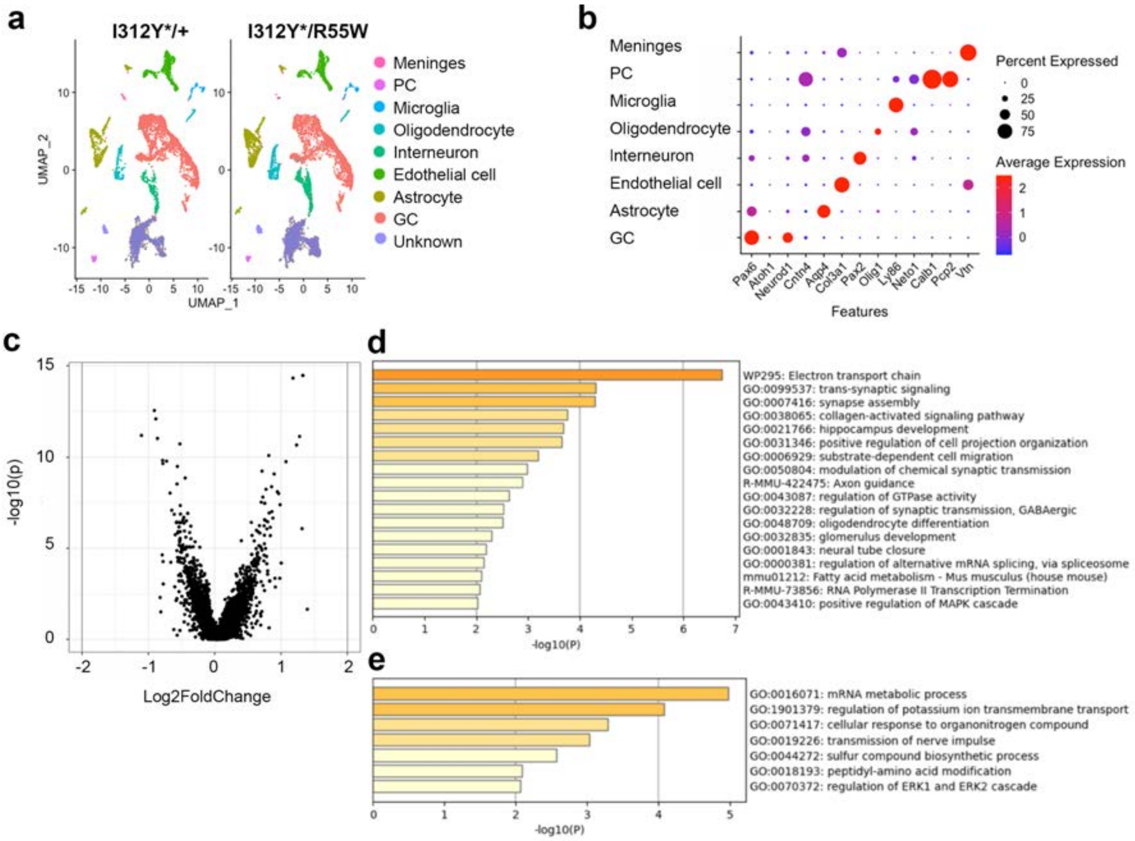


**Supplementary Figure 6. snRNA-seq analysis for whole cerebellum of I312Y*/+ and I312Y*/R55W**

(a) Two-dimensional UMAP distribution of snRNA-seq of both genotypes. UMAP divided into 9 clustered by R-analysis (resolution=0.02). (b) Identification of cell clusters based on the expression of known genes. (c) Volcanoplot of PC cluster of snRNA-seq. There are 167 up regulated genes and 58 down regulated genes (log fold change≧ 0.5 or ≦ -0.5). Enriched ontology clustered of 167 up regulated genes (d) and 58 down regulated genes (e) by Metascape.

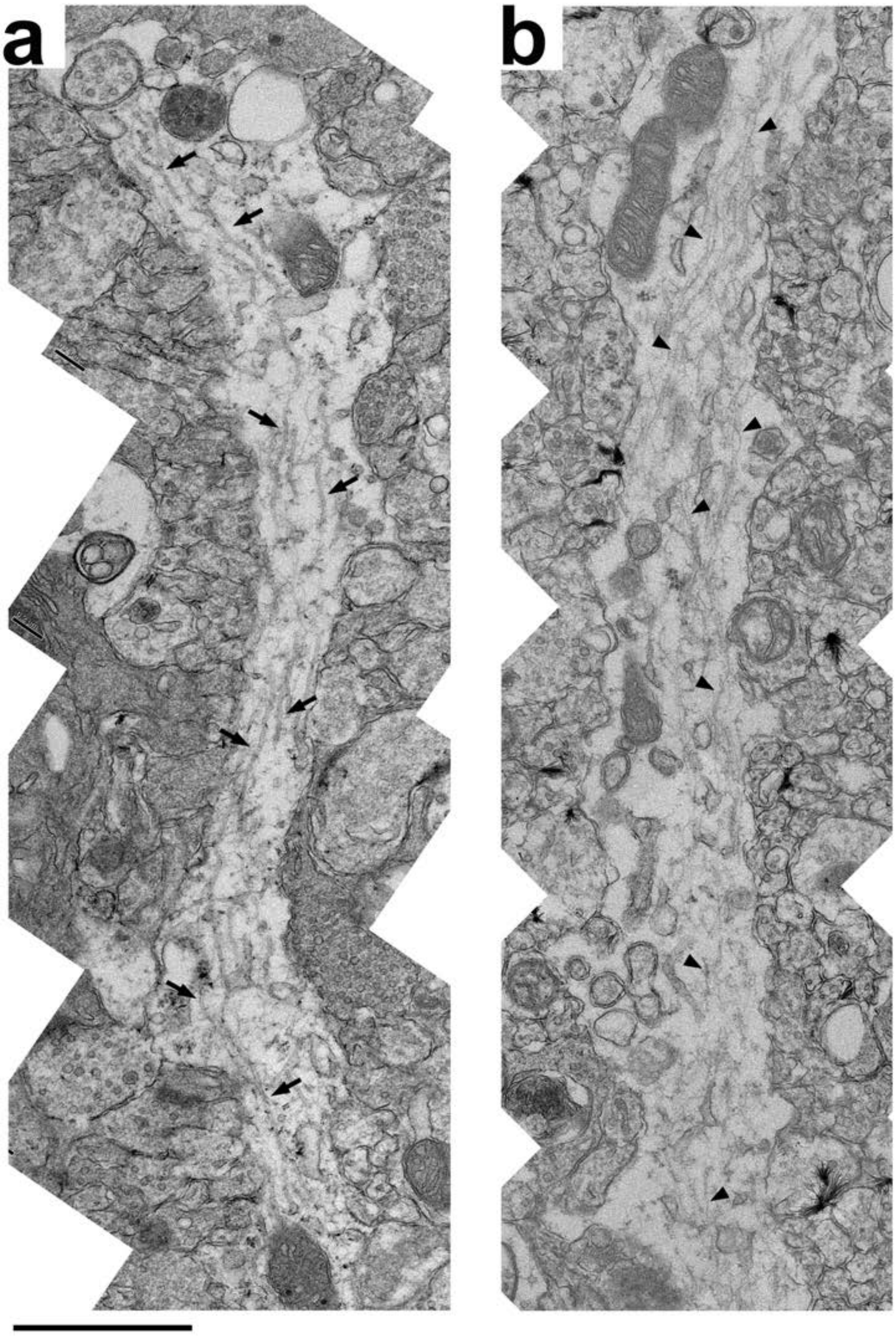


**Supplementary Figure 7. Electron micrographs of proximal dendritic domain in PCs.**

Electron micrographs of proximal dendritic domain in PC of control (I312Y*/+) (a) and mutant (I312Y*/R55W) (b) mice at the age of P10. Black arrows and triangles show microtubules. Scale bar, 1 μm

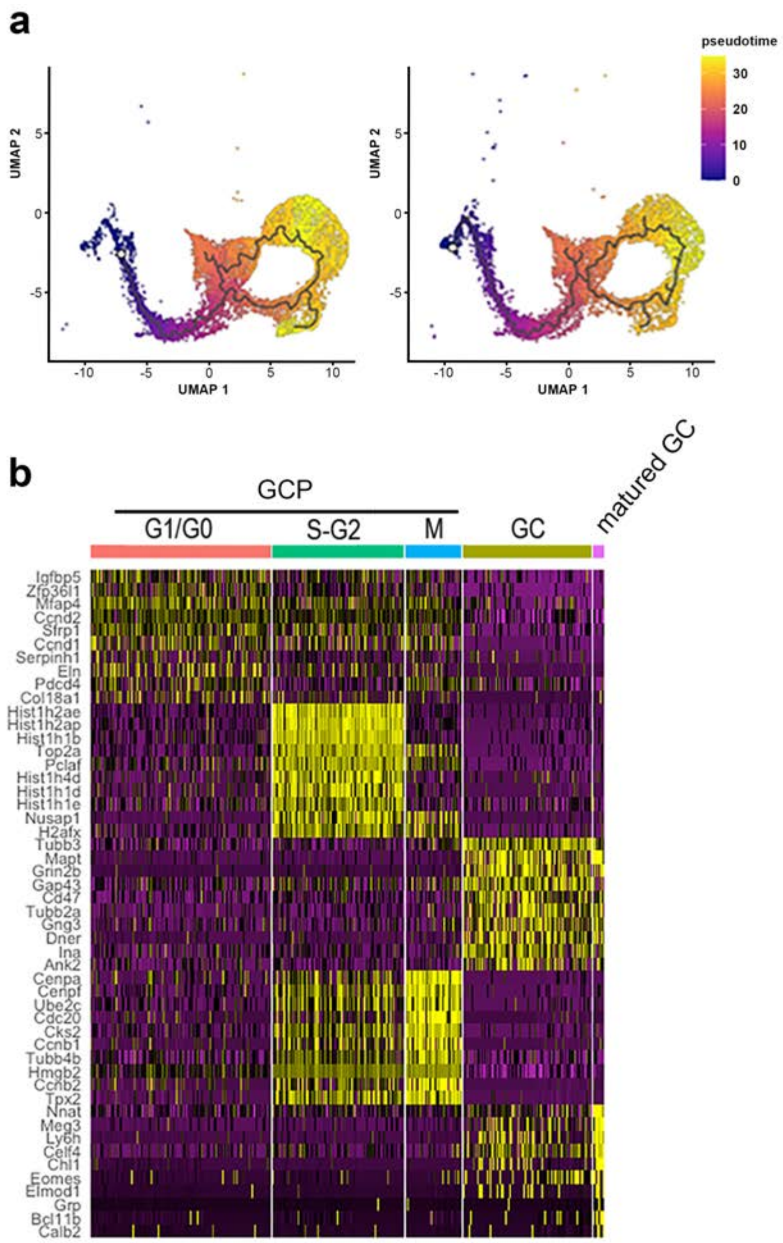


**Supplementary Figure 8. Subcluster analysis for granule cells (GCs) and granule cell progenitors (GCPs)**

(a) Two-dimensional UMAP distribution of cell clusters in the GC by pseudo time for both I312Y*/+ (left) and I312Y*/R55W (right). (b) Heatmap of specific gene expression in GCs and GCPs grouped by differentiation trajectory (Figure 6c).

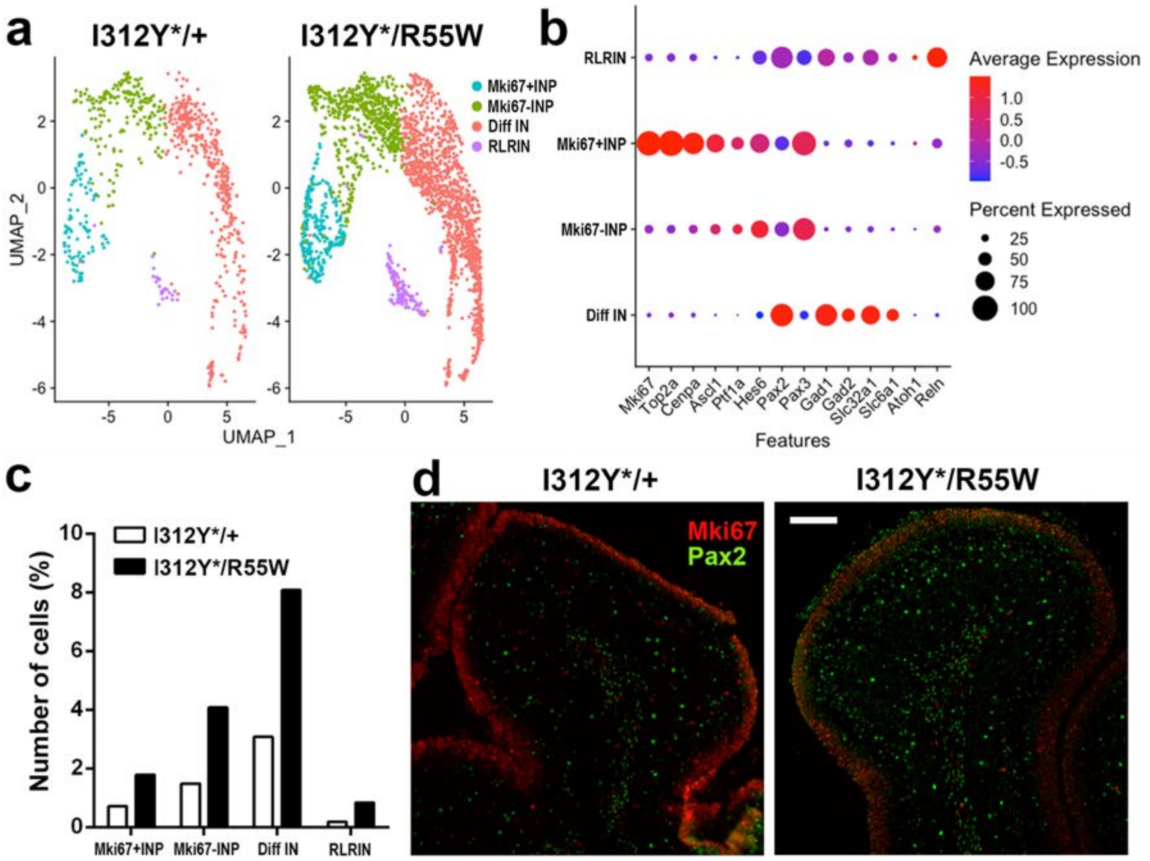


**Supplementary Figure 9. Change of inhibitory interneurons (INs) in cerebella**

(a) Two-dimensional UMAP distribution of INs clusters of both genotypes.

UMAP divided into 4 clustered by R-analysis (resolution=0.1). (b) Dotplot showing known genes for proliferation (*Mki67, Top2a, Cenpa*), differentiation (*Asd1, Ptf1a, Hes6, Pax2, Pax3, Gad1*) and maturation (*Gad2, Slc32a1, Slc6a1, Atoh1, Reln*). (c) Percentage of cell number in INs subclusters. (d) Immunostaining for Mki67 and Pax2 for I312Y*/+ and I312Y*/R55W. Pax2 positive INs were increased in molecular layer and white matters on cerebellum section from I312Y*/R55W. Scale bar, 100 um
